# Association of Neutrophil-to-Lymphocyte Ratio and Systemic Immune-Inflammation Index With Mortality in Patients With Pericarditis: A Retrospective Dual-Cohort Study Using Two Independent Databases

**DOI:** 10.64898/2026.06.25.26356550

**Authors:** Lingyu Mi, Ishan Lakhani, Wing Tak Wong, Gary Tse, Fang Fang

## Abstract

**Background:** Risk stratification in pericarditis relies mainly on clinical presentation, suspected etiology, imaging findings, and conventional inflammatory biomarkers. Whether complete blood count–derived inflammatory indices are associated with mortality in pericarditis and reproducible across independent real-world datasets remains unclear.

**Methods:** We conducted a retrospective dual-cohort study of hospitalized adults with pericarditis using a Hong Kong cohort from the Clinical Data Analysis and Reporting System (CDARS) as the primary analysis cohort and the Medical Information Mart for Intensive Care IV (MIMIC-IV) cohort as an independent reproducibility cohort. Baseline neutrophil-to-lymphocyte ratio (NLR) and systemic immune-inflammation index (SII) were analyzed as continuous variables and cohort-specific tertiles. The primary outcome was long-term all-cause mortality in the Hong Kong cohort. Secondary and reproducibility outcomes included 90-day mortality in the Hong Kong cohort and 30-day, 90-day, and observable follow-up mortality in MIMIC-IV. Cox models were adjusted for age, sex, renal disease, diabetes mellitus, hypertension, ischemic heart disease, and malignancy.

**Results:** Among 504 patients in the Hong Kong cohort and 464 patients in MIMIC-IV, all-cause mortality occurred in 241 and 113 patients during cohort-specific follow-up, respectively. In the Hong Kong cohort, higher NLR was associated with long-term all-cause mortality after full adjustment. Compared with NLR tertile 1, the adjusted hazard ratio was 1.60 for tertile 3. Higher SII was also associated with long-term mortality, with an adjusted hazard ratio of 1.55 for tertile 3 versus tertile 1. NLR and SII showed directionally consistent associations with 90-day mortality in the Hong Kong cohort and with 30-day, 90-day, and observable follow-up mortality in MIMIC-IV. Sensitivity analyses yielded broadly consistent findings.

**Conclusions:** In two independent real-world cohorts of hospitalized patients with pericarditis, higher baseline NLR and SII were associated with increased all-cause mortality, with NLR showing the more consistent prognostic signal. These complete blood count-derived indices may provide simple adjunctive information for mortality risk stratification, although prospective validation is needed before incorporation into formal management algorithms.

## Introduction

Pericarditis is part of a heterogeneous spectrum of pericardial diseases and has a variable clinical course^1^. Although most patients have a benign course with appropriate treatment, a subset develops recurrent or prolonged disease, hospitalization, or life-threatening complications such as cardiac tamponade and constrictive pericarditis^2^. Accurate risk stratification is therefore important for identifying patients who may require closer surveillance or more intensive treatment^1,2^. In routine practice, risk assessment relies mainly on clinical presentation, suspected etiology, imaging findings, and inflammatory biomarkers such as C-reactive protein^2,3^. However, simple and widely available blood-based indicators for mortality risk stratification in patients with pericarditis remain insufficiently established.

Systemic inflammation is central to the pathophysiology and clinical expression of pericarditis^1,4^. In real-world patients, inflammatory activation may reflect not only local pericardial injury but also infection, immune dysregulation, malignancy, renal dysfunction, and cardiovascular vulnerability^4^. Although C-reactive protein is clinically useful for diagnosis, assessment of disease activity, and treatment monitoring, it may not fully capture the broader inflammatory and immune status associated with adverse outcomes in pericarditis^2,4^. Therefore, inflammatory indices derived from complete blood count parameters may provide a simple and widely available approach to complement conventional risk assessment^4,5^.

The neutrophil-to-lymphocyte ratio (NLR) reflects the balance between neutrophil-mediated innate inflammatory activation and lymphocyte-related adaptive immune status^6^. The systemic immune-inflammation index (SII), calculated from platelet, neutrophil, and lymphocyte counts, further integrates platelet-related thromboinflammatory information^7^. Both indices have been associated with adverse cardiovascular outcomes and mortality in several cardiovascular and inflammatory conditions^7,8^. However, whether these complete blood count-derived indices are associated with mortality in patients with pericarditis, and whether such associations are reproducible across independent real-world datasets, remains unclear.

Previous studies of inflammatory biomarkers in pericarditis have largely focused on disease activity, recurrence, inflammatory phenotypes, or short-term complications, whereas evidence regarding mortality outcomes remains limited^9–12^. Moreover, most available studies have been conducted in single cohorts, with limited assessment of reproducibility across independent real-world datasets. Therefore, studies evaluating the prognostic relevance and cross-cohort reproducibility of complete blood count-derived inflammatory indices may help clarify their potential role as pragmatic risk stratification markers in pericarditis.

In this retrospective dual-cohort study, we evaluated whether baseline NLR and SII were associated with all-cause mortality in patients with pericarditis using two independent real-world databases. The study was designed to address three clinically relevant gaps: the limited evidence on complete blood count-derived inflammatory indices for mortality risk stratification in pericarditis, the lack of evaluation across both short- and long-term mortality horizons, and the limited assessment of reproducibility across independent electronic health record datasets. The Hong Kong cohort served as the primary analysis cohort, with long-term all-cause mortality as the primary outcome and 90-day all-cause mortality as a secondary outcome. The MIMIC-IV cohort was used for reproducibility analyses of 30-day, 90-day, and observable follow-up all-cause mortality. We hypothesized that higher baseline NLR and SII would identify patients at increased mortality risk and that these associations would be directionally reproducible in an independent cohort.

## Methods

### Study design, data sources, and population

We conducted a retrospective dual-cohort study of adult patients with pericarditis using two independent electronic health record datasets. The Hong Kong cohort was used as the primary analysis cohort, and the Medical Information Mart for Intensive Care IV (MIMIC-IV) cohort was used as an independent reproducibility cohort.

The Hong Kong cohort was derived from a previously conducted retrospective cohort study of adult patients hospitalized with a clinical diagnosis with an index date of pericarditis between January 1, 2005, and December 31, 2019, at a single tertiary centre in Hong Kong. This study utilised publicly available, deidentified data that arose directly from clinical data collected as part of GT’s doctoral thesis towards the Doctor of Medicine Degree conducted at the Division of Graduate Studies, The Chinese University of Hong Kong accompanied by ethics approvals from the Joint Chinese University of Hong Kong-New Territories East Cluster Clinical Research Ethics Committee, with approval numbers 2018.309, 2018.643, 2019.338, 2019.361. The requirement for informed consent was waived because of the retrospective design and use of deidentified data. It is a subanalysis of a published study that has been shared on the preprint server MedRxiv (https://www.medrxiv.org/content/10.1101/2021.11.07.21266025v1). The analyses of deidentified Hong Kong data were completed in June 2023, within the registration period of G.T at the Chinese University of Hong Kong. Written approval from the Doctor of Medicine Subcommittee for the use of data was provided to G.T. in July 2025. An anonymised version of the compiled database without personal details or the underlying raw data has been made available on ProQuest as part of the M.D. thesis (https://www.proquest.com/docview/3252768154/278E578A36174385PQ/2?sourcetype=Dissertations%20&%20Theses). The submission to ProQuest was managed and approved by the Graduate School of the Chinese University of Hong Kong as part of the graduation requirements of G.T.. The Clinical Data Analysis and Reporting System is a territory-wide database that centralizes patient information from local hospitals and associated ambulatory and outpatient facilities, including diagnoses, laboratory results, clinical characteristics, and treatment records. This system has been used extensively by local research teams ^13–15^, to conduct cross-cluster clinical research studies using territory-wide data ^16^.

MIMIC-IV version 2.2 is a publicly available, deidentified electronic health record database derived from patients admitted to Beth Israel Deaconess Medical Center in Boston, Massachusetts, between 2008 and 2019^17^. Access was granted after completion of the required data use training. Use of the database was approved by the institutional review boards of the Massachusetts Institute of Technology and Beth Israel Deaconess Medical Center, with a waiver of informed consent. Data were extracted using Structured Query Language in a PostgreSQL environment. Pericarditis was identified using International Classification of Diseases, Ninth and Tenth Revision diagnostic codes.

In both cohorts, adult patients aged 18 years or older with pericarditis were eligible. For patients with multiple qualifying hospitalizations, only the index admission was retained. Patients were excluded if neutrophil, lymphocyte, or platelet measurements were unavailable, because these variables were required to calculate the neutrophil-to-lymphocyte ratio (NLR) and systemic immune-inflammation index (SII). Patients without survival outcome data were also excluded. In the Hong Kong cohort, baseline laboratory measurements were defined as the first available results obtained at admission or during the early hospitalization period of the index admission. In the MIMIC-IV cohort, eligible laboratory measurements were extracted from 6 hours before to 24 hours after hospital admission whenever available. Among 874 patients initially identified in the Hong Kong cohort, 504 had available NLR and SII measurements and were included in the main analyses. Among 661 patients identified in MIMIC-IV, 464 had available NLR and SII measurements and were included in the reproducibility analyses.

### Exposure variables and outcomes

The primary exposure was baseline NLR, calculated as the neutrophil count divided by the lymphocyte count. The secondary exposure was SII, calculated as platelet count multiplied by neutrophil count and divided by lymphocyte count. Both indices were analyzed as continuous variables and as cohort-specific tertiles. Tertiles were defined separately within each cohort to account for differences in case mix and biomarker distributions. In the Hong Kong cohort, the upper cut-off values for NLR tertile 1 and tertile 2 were 2.80 and 6.57, respectively, and the corresponding cut-off values for SII were 569.99 and 1460.21. In MIMIC-IV, the upper cut-off values for NLR tertile 1 and tertile 2 were 4.08 and 8.68, respectively, and the corresponding cut-off values for SII were 1002.53 and 2472.08. SII was modeled per 100-unit increase in continuous analyses.

The primary outcome was long-term all-cause mortality in the Hong Kong cohort during cohort-specific follow-up. Secondary mortality analysis in the Hong Kong cohort included 90-day all-cause mortality. In the Hong Kong cohort, vital status and dates of death were ascertained from the Clinical Data Analysis and Reporting System, which integrates electronic health records from local hospitals and associated ambulatory and outpatient facilities. In MIMIC-IV, reproducibility analyses were performed for 30-day all-cause mortality, 90-day all-cause mortality, and all-cause mortality during observable follow-up. Vital status and dates of death in MIMIC-IV were obtained from the deidentified date-of-death information available in the database and were used to derive 30-day, 90-day, and observable follow-up all-cause mortality. Cohort-specific follow-up referred to long-term electronic health record follow-up in the Hong Kong cohort and observable follow-up available in MIMIC-IV. Follow-up duration was defined as the time from index admission to death or the last available follow-up record.

### Covariates

Covariates were selected before analysis according to clinical relevance, availability in both cohorts, event numbers, and their potential associations with inflammatory biomarkers and mortality. To improve comparability between the primary and reproducibility analyses, adjusted models in both cohorts used a common covariate set, including age, sex, renal disease, diabetes mellitus, hypertension, ischemic heart disease, and malignancy. Variables with substantial missingness or limited availability across cohorts, including C-reactive protein and other inflammatory or cardiac injury biomarkers, were not included in the primary multivariable models.

### Statistical analysis

Continuous variables were presented as medians with interquartile ranges, and categorical variables were presented as counts and percentages. Continuous variables were compared using the Wilcoxon rank-sum test, and categorical variables were compared using the chi-square test or Fisher exact test, as appropriate.

Missing data were handled using a complete-case approach. Patients with missing neutrophil, lymphocyte, or platelet counts were excluded because NLR and SII could not be calculated, and no imputation was performed for exposure variables. Missingness of key variables was summarized in **Supplementary Table 3**.

Baseline characteristics were summarized separately for the Hong Kong and MIMIC-IV cohorts and were further summarized according to NLR tertiles in the Hong Kong cohort. P values for trend across NLR tertiles were calculated using a rank-based linear trend test for continuous variables and the Cochran-Armitage trend test for categorical variables.

Kaplan-Meier cumulative mortality curves were generated according to NLR and SII tertiles, and differences between groups were assessed using log-rank tests. Cox proportional hazards regression models were used to evaluate the associations of NLR and SII with all-cause mortality outcomes. For categorical analyses, the lowest tertile served as the reference group. Linear trends across tertiles were assessed by modeling tertiles as an ordinal variable.

In the Hong Kong cohort, the primary analysis examined long-term all-cause mortality. Sequential Cox models were constructed: Model 1 was unadjusted; Model 2 was adjusted for age and sex; and Model 3 was adjusted for age, sex, renal disease, diabetes mellitus, hypertension, ischemic heart disease, and malignancy. Secondary mortality analysis in the Hong Kong cohort evaluated 90-day all-cause mortality using the same covariate set as Model 3. Reproducibility analyses in MIMIC-IV evaluated 30-day, 90-day, and observable follow-up all-cause mortality using the same covariate set. For 30-day and 90-day mortality analyses, follow-up was administratively censored at 30 and 90 days, respectively. The proportional hazards assumption was assessed using Schoenfeld residuals.

Restricted cubic spline analyses were performed in the Hong Kong cohort to examine dose-response associations of NLR and SII with long-term all-cause mortality. Spline models were adjusted for the same covariates as Model 3 and were plotted from the 2.5th to 97.5th percentiles to reduce the influence of extreme values.

Sensitivity analyses were conducted to assess the robustness of the findings. In the Hong Kong cohort, these included analyses with follow-up truncated at 5 years to evaluate the robustness of the long-term Cox models in relation to possible time-varying effects, analyses of individual blood cell components restricted to the main analytical cohort, and extreme-value analyses by excluding patients above the 99th percentile of the corresponding biomarker or by winsorizing biomarker values at the 1st and 99th percentiles. Because the numbers of early mortality events were limited, additional parsimonious multivariable Cox models adjusted for age, sex, renal disease, and malignancy were performed for short-term and observable follow-up mortality outcomes to reduce the risk of overfitting. The incremental prognostic value of NLR and SII was assessed by comparing a clinical model with models additionally incorporating NLR, SII, or both indices. Model performance was evaluated using Harrell’s C-index, time-dependent areas under the receiver operating characteristic curves, integrated discrimination improvement, and continuous net reclassification improvement.

All statistical analyses were conducted using R software (version 4.6.0; R Foundation for Statistical Computing, Vienna, Austria). All statistical tests were two-sided, and a P value <0.05 was considered statistically significant.

## Results

### Study population and baseline characteristics

The study flowchart is shown in **Figure 1**. Among 874 patients with pericarditis identified in the Hong Kong cohort, 504 had available NLR and SII measurements and were included in the main analyses. Among 661 patients identified in the MIMIC-IV cohort, 464 had available NLR and SII measurements and were included in the reproducibility analyses.

**Figure 1.**
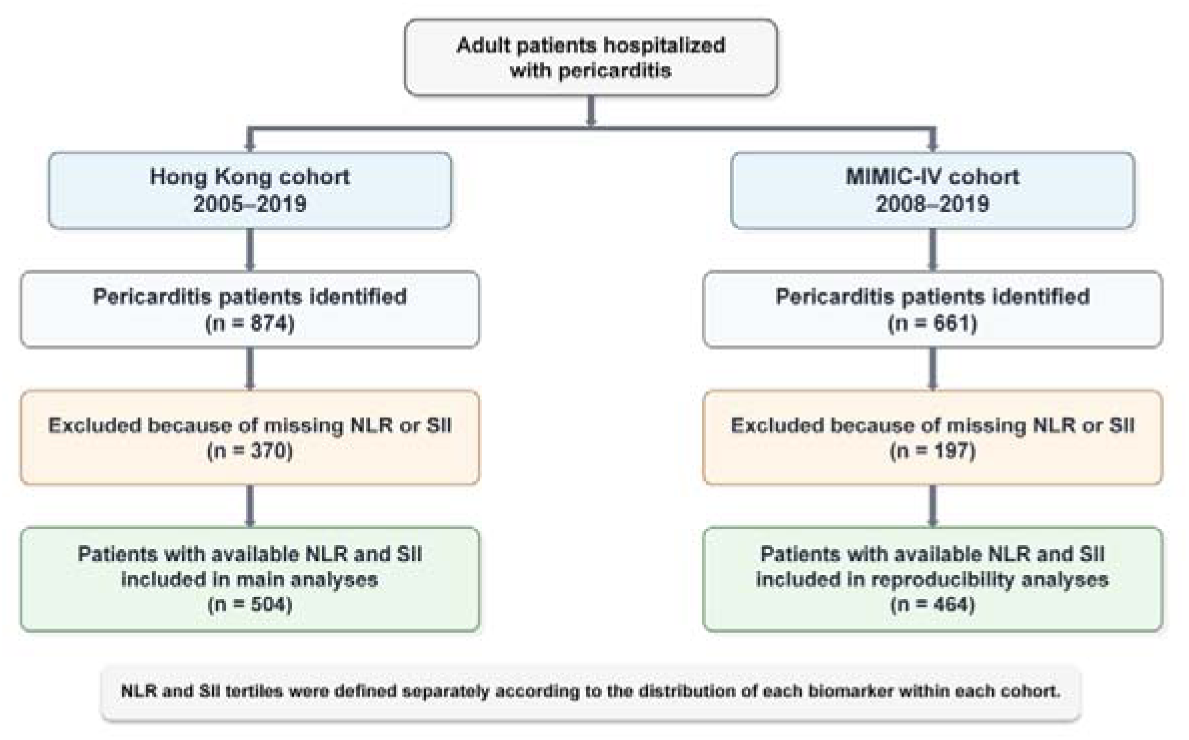
Study flowchart. Flowchart of patient selection in the Hong Kong and MIMIC-IV cohorts. The Hong Kong cohort included adult patients hospitalized with pericarditis between 2005 and 2019, and the MIMIC-IV cohort included adult patients hospitalized with pericarditis between 2008 and 2019. Patients with available NLR and SII were included in the main analysis of the Hong Kong cohort or the reproducibility analysis of the MIMIC-IV cohort. NLR and SII tertiles were defined separately according to the distribution of each biomarker within each cohort. **Abbreviations:** NLR, neutrophil-to-lymphocyte ratio; SII, systemic immune-inflammation index.

Baseline characteristics of the two cohorts are presented in **Table 1**. The median age was 60.70 years in the Hong Kong cohort and 59.00 years in the MIMIC-IV cohort. Male patients accounted for 52.2% and 51.7% of the two cohorts, respectively. Compared with the Hong Kong cohort, the MIMIC-IV cohort had higher proportions of renal disease, diabetes mellitus, hypertension, acute myocardial infarction, chronic obstructive pulmonary disease, ischemic heart disease, and peripheral vascular disease. The median NLR was 4.08 in the Hong Kong cohort and 5.81 in the MIMIC-IV cohort, and the median SII was 826.49 and 1627.06, respectively. During cohort-specific follow-up, all-cause mortality occurred in 241 patients in the Hong Kong cohort and 113 patients in the MIMIC-IV cohort. The median follow-up duration was 1980.00 days and 175.00 days, respectively.

**Table 1.** Baseline characteristics of patients with available NLR and SII in the Hong Kong and MIMIC-IV cohorts.

| Characteristic | Hong Kong<br>(n=504) | MIMIC-IV<br>(n=464) | P value |
| --- | --- | --- | --- |
| **Demographics** |  |  |  |
| Age, years | 60.70 (50.19, 71.78) | 59.00 (46.00, 69.00) | 0.051 |
| Male sex, n (%) | 263 (52.2%) | 240 (51.7%) | 0.938 |
| **Comorbidities** |  |  |  |
| History of VT/VF | 4 (0.8%) | 22 (4.7%) | <0.001 |
| Cerebrovascular disease | 17 (3.4%) | 29 (6.2%) | 0.051 |
| Renal disease | 33 (6.5%) | 173 (37.3%) | <0.001 |
| Diabetes mellitus | 14 (2.8%) | 108 (23.3%) | <0.001 |
| Hypertension | 97 (19.2%) | 194 (41.8%) | <0.001 |
| Acute myocardial infarction | 11 (2.2%) | 53 (11.4%) | <0.001 |
| COPD | 5 (1.0%) | 55 (11.9%) | <0.001 |
| Ischemic heart disease | 55 (10.9%) | 108 (23.3%) | <0.001 |
| Peripheral vascular disease | 6 (1.2%) | 36 (7.8%) | <0.001 |
| Malignancy | 101 (20.0%) | 90 (19.4%) | 0.865 |
| Metastatic solid tumor | 47 (9.3%) | 42 (9.1%) | 0.971 |
| **Laboratory indices** |  |  |  |
| White blood cell count, $\times 10^9/L$ | 7.05 (5.50, 9.03) | 9.90 (7.50, 13.40) | <0.001 |
| Platelet count, $\times 10^9/L$ | 210.00 (163.00, 274.25) | 258.00 (196.75, 345.50) | <0.001 |
| NLR | 4.08 (2.43, 8.56) | 5.81 (3.36, 10.60) | <0.001 |
| SII | 826.49 (468.16, 1911.88) | 1627.06 (782.59, 2992.41) | <0.001 |
| Creatinine, $\mu\text{mol/L}$ | 85.00 (66.00, 119.00) | 88.40 (70.72, 106.08) | 0.883 |
| Glucose, mmol/L | 6.39 (5.41, 8.20) | 6.33 (5.44, 7.56) | 0.748 |
| **Outcomes and follow-up** |  |  |  |
| 90-day all-cause mortality, n (%) | 33 (6.5%) | 41 (8.8%) | 0.223 |
| All-cause mortality during cohort-specific follow-up, n (%) | 241 (47.8%) | 113 (24.4%) | <0.001 |
| Follow-up duration, days | 1980.00 (967.75, 3172.25) | 175.00 (40.00, 487.00) | <0.001 |

Continuous variables are presented as median (interquartile range), and categorical variables are presented as n (%). P values were calculated using the Wilcoxon rank-sum test for continuous variables and the chi-square test or Fisher's exact test for categorical variables, as appropriate. Creatinine and glucose were harmonized to $\mu\text{mol/L}$ and $\text{mmol/L}$ , respectively. Cohort-specific follow-up refers to long-term electronic health record follow-up in the Hong Kong cohort and observable follow-up available in the MIMIC-IV database. For the Hong Kong cohort, the primary outcome was long-term all-cause mortality during cohort-specific follow-up. For the MIMIC-IV cohort, all-cause mortality during cohort-specific follow-up was used as part of the reproducibility analyses. P values for outcomes and follow-up duration are descriptive because follow-up windows differed between cohorts.
**Abbreviations:** VT/VF, ventricular tachycardia/ventricular fibrillation; COPD, chronic obstructive pulmonary disease; NLR, neutrophil-to-lymphocyte ratio; SII, systemic immune-inflammation index.

Characteristics of patients with and without available NLR and SII measurements are shown in **Supplementary Table 1** for the Hong Kong cohort and **Supplementary Table 2** for the MIMIC-IV cohort. Missingness of key variables in both cohorts is summarized in **Supplementary Table 3**.

### Baseline characteristics according to NLR tertiles in the Hong Kong cohort

Baseline characteristics and clinical outcomes according to NLR tertiles in the Hong Kong cohort are shown in **Table 2**. Patients were divided into three equal tertiles, with 168 patients in each group. The upper cut-off values for NLR tertile 1 and tertile 2 were 2.80 and 6.57, respectively. Across increasing NLR tertiles, white blood cell count, neutrophil count, platelet count, and SII increased, whereas lymphocyte count decreased. Age, renal disease, diabetes mellitus, hypertension, ischemic heart disease, and malignancy did not differ significantly across NLR tertiles.

**Table 2.**
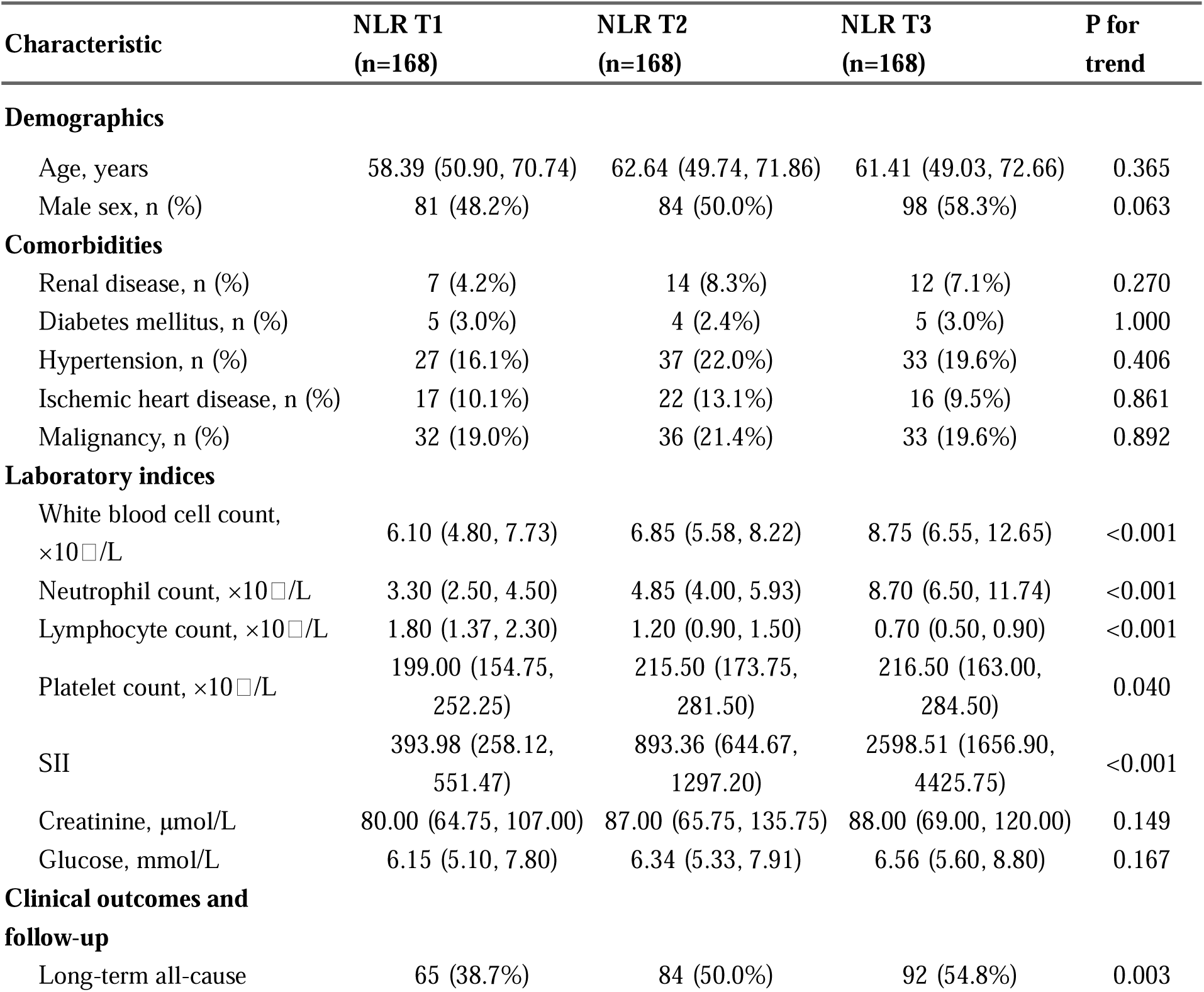
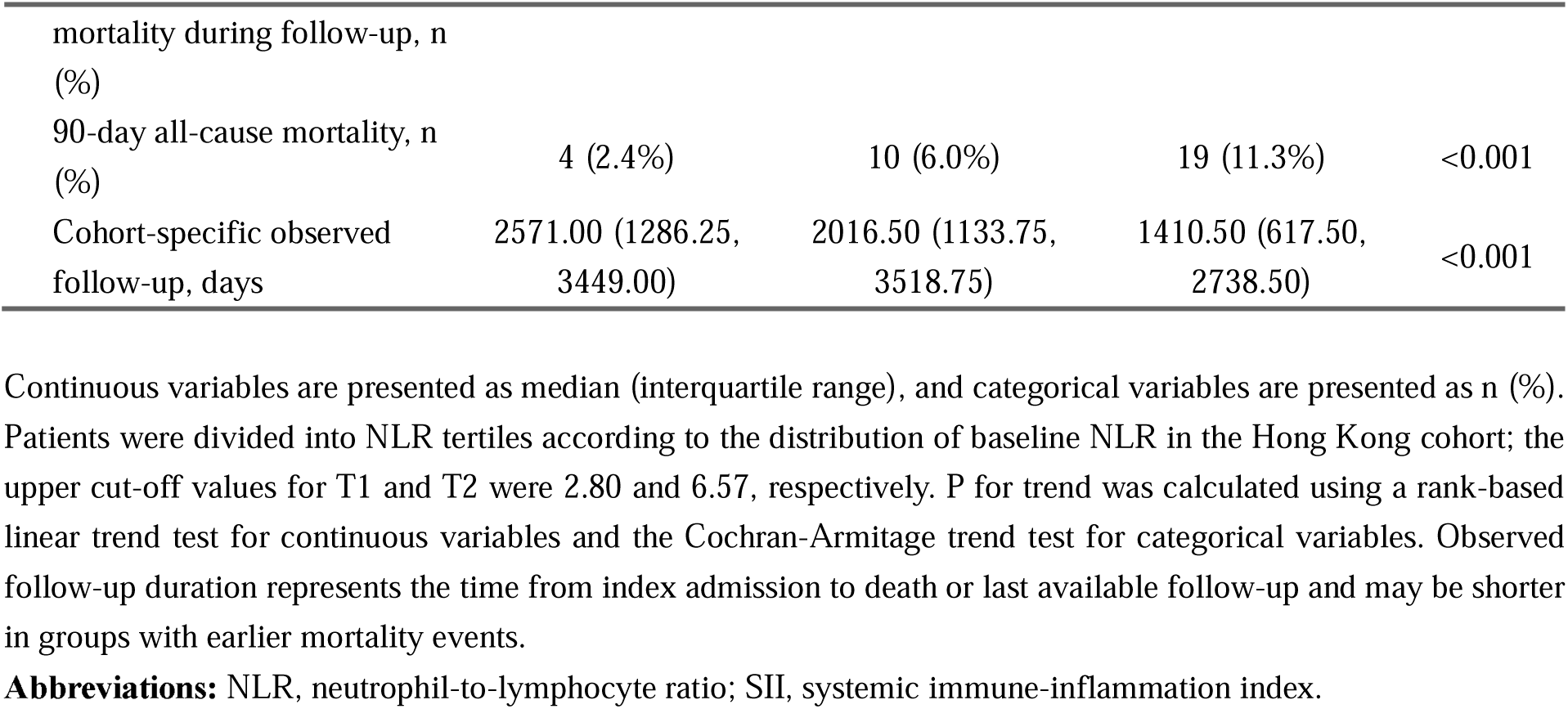
Baseline characteristics and clinical outcomes according to NLR tertiles in the Hong Kong cohort.

Long-term all-cause mortality occurred in 65 patients in NLR tertile 1, 84 patients in tertile 2, and 92 patients in tertile 3. The corresponding numbers of 90-day all-cause deaths were 4, 10, and 19, respectively. Observed follow-up duration decreased across increasing NLR tertiles.

### NLR, SII, and long-term all-cause mortality in the Hong Kong cohort

Cox proportional hazards regression analyses for long-term all-cause mortality in the Hong Kong cohort are presented in **Table 3**. In unadjusted analyses, NLR was associated with long-term all-cause mortality as a continuous variable and across tertiles. Compared with NLR tertile 1, the unadjusted HRs were 1.39 for tertile 2 and 1.87 for tertile 3. After adjustment for age and sex, the HR for tertile 3 versus tertile 1 was 1.61. In the fully adjusted model including age, sex, renal disease, diabetes mellitus, hypertension, ischemic heart disease, and malignancy, NLR was associated with long-term all-cause mortality as a continuous variable. Compared with NLR tertile 1, the adjusted HR was 1.60 for tertile 3, with a significant trend across tertiles (P for trend = 0.006).

**Table 3.**
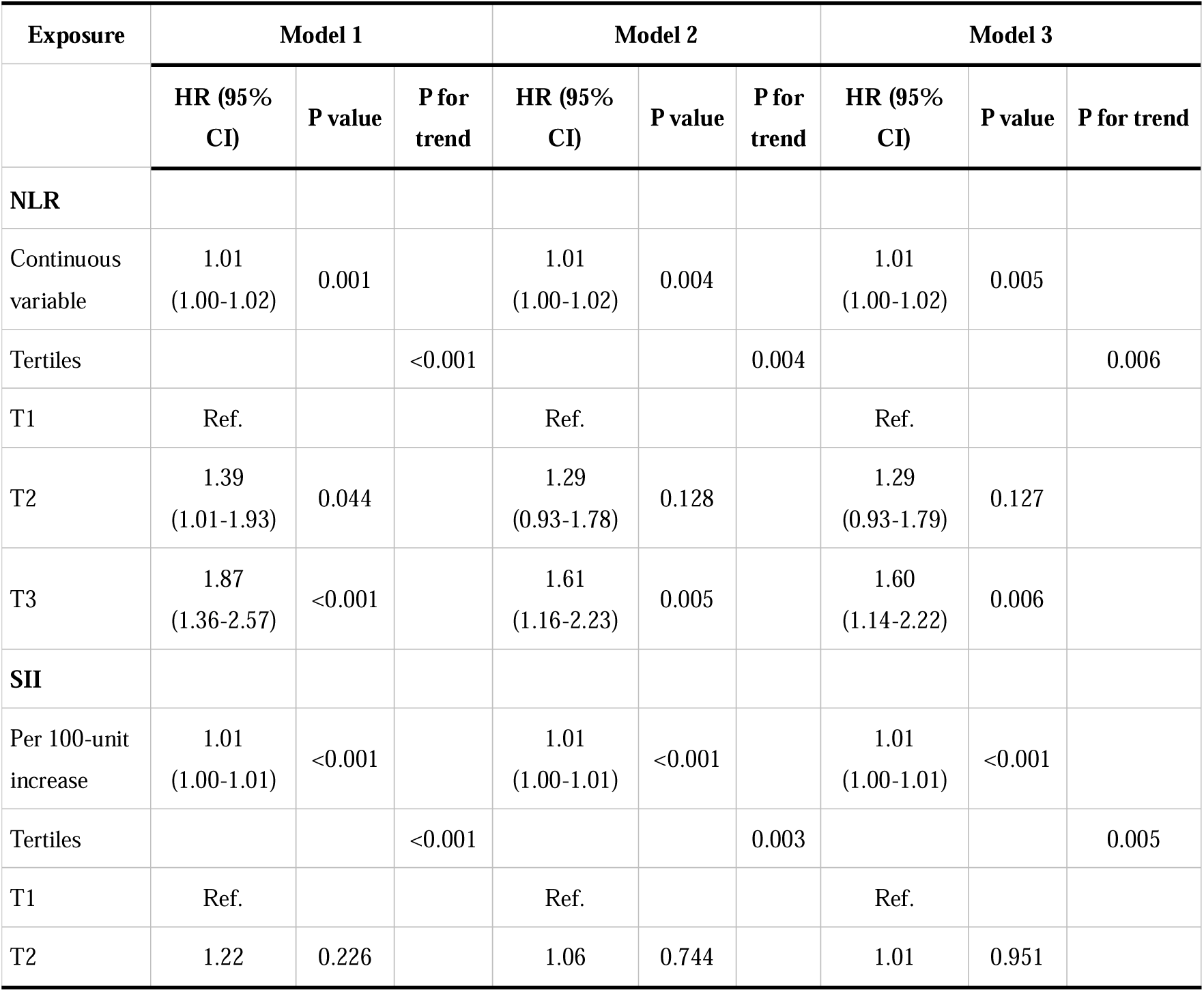
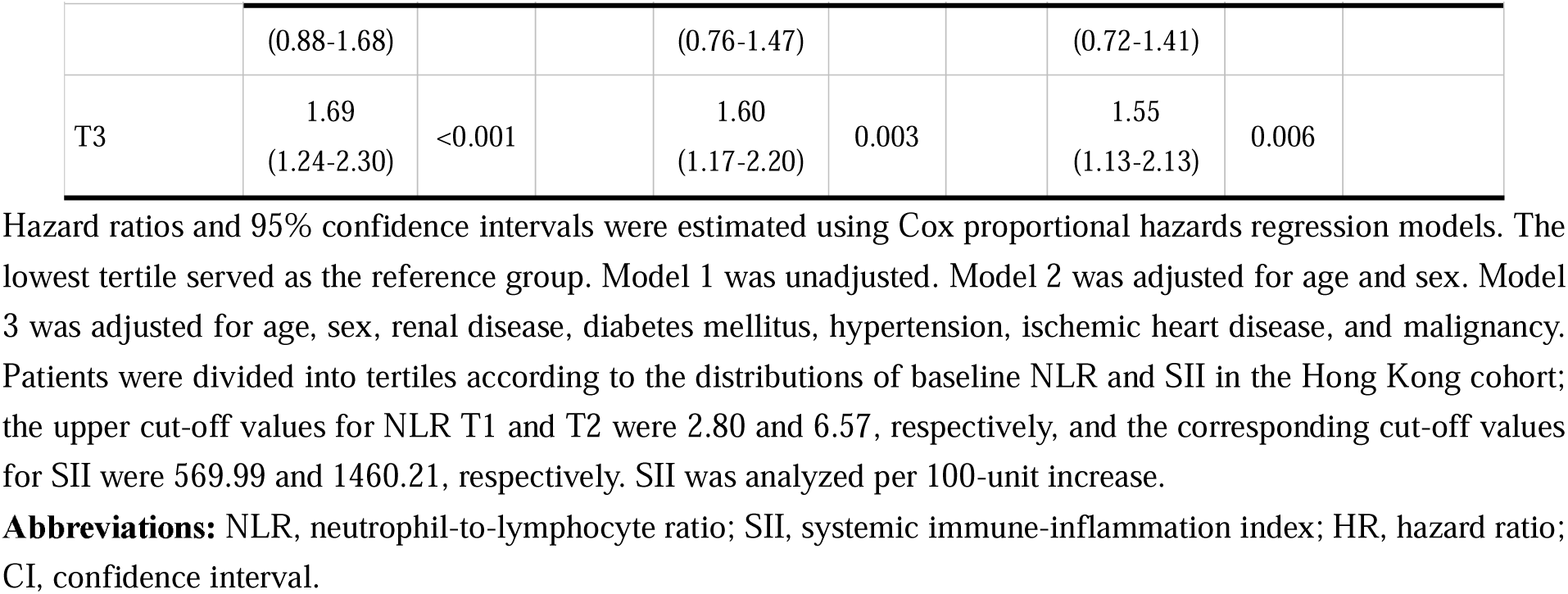
Cox proportional hazards regression analyses for long-term all-cause mortality according to NLR and SII in the Hong Kong cohort.

SII was also analyzed in relation to long-term all-cause mortality (**Table 3**). In the fully adjusted model, SII was associated with long-term all-cause mortality when modeled per 100-unit increase. Compared with SII tertile 1, the adjusted HR was 1.55 for tertile 3, with a significant trend across tertiles (P for trend = 0.005).

Kaplan-Meier cumulative mortality curves according to NLR tertiles are shown in **Figure 2**. In the Hong Kong cohort, cumulative long-term all-cause mortality differed across NLR tertiles (log-rank P < 0.001). Cumulative 90-day all-cause mortality also differed across NLR tertiles (log-rank P = 0.004). Kaplan-Meier curves according to SII tertiles are shown in **Supplementary Figure 1**.

**Figure 2.**
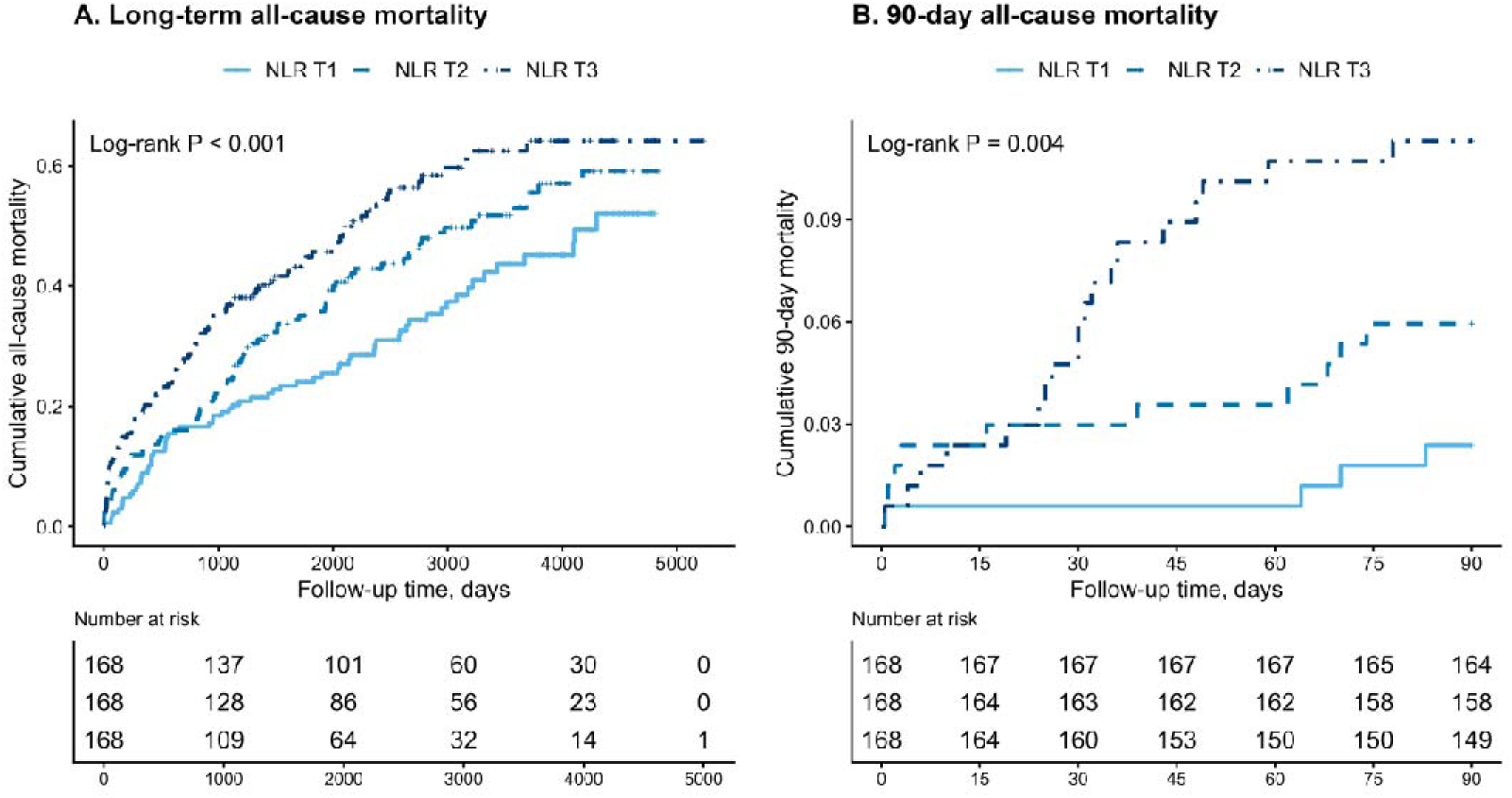
Kaplan-Meier cumulative mortality curves according to NLR tertiles in the Hong Kong cohort. Kaplan-Meier cumulative mortality curves for patients stratified by baseline NLR tertiles in the Hong Kong cohort. (A) Long-term all-cause mortality. (B) 90-day all-cause mortality. Patients were divided into NLR tertiles according to the distribution of baseline NLR; the upper cut-off values for NLR T1 and T2 were 2.80 and 6.57, respectively. Log-rank tests were used to compare cumulative mortality across NLR tertiles. **Abbreviations:** NLR, neutrophil-to-lymphocyte ratio.

Restricted cubic spline analyses of NLR, SII, and long-term all-cause mortality in the Hong Kong cohort are shown in **Figure 3**. For NLR, the overall association with long-term all-cause mortality was significant, whereas the test for nonlinearity was not significant. Similar findings were observed for SII.

**Figure 3.**
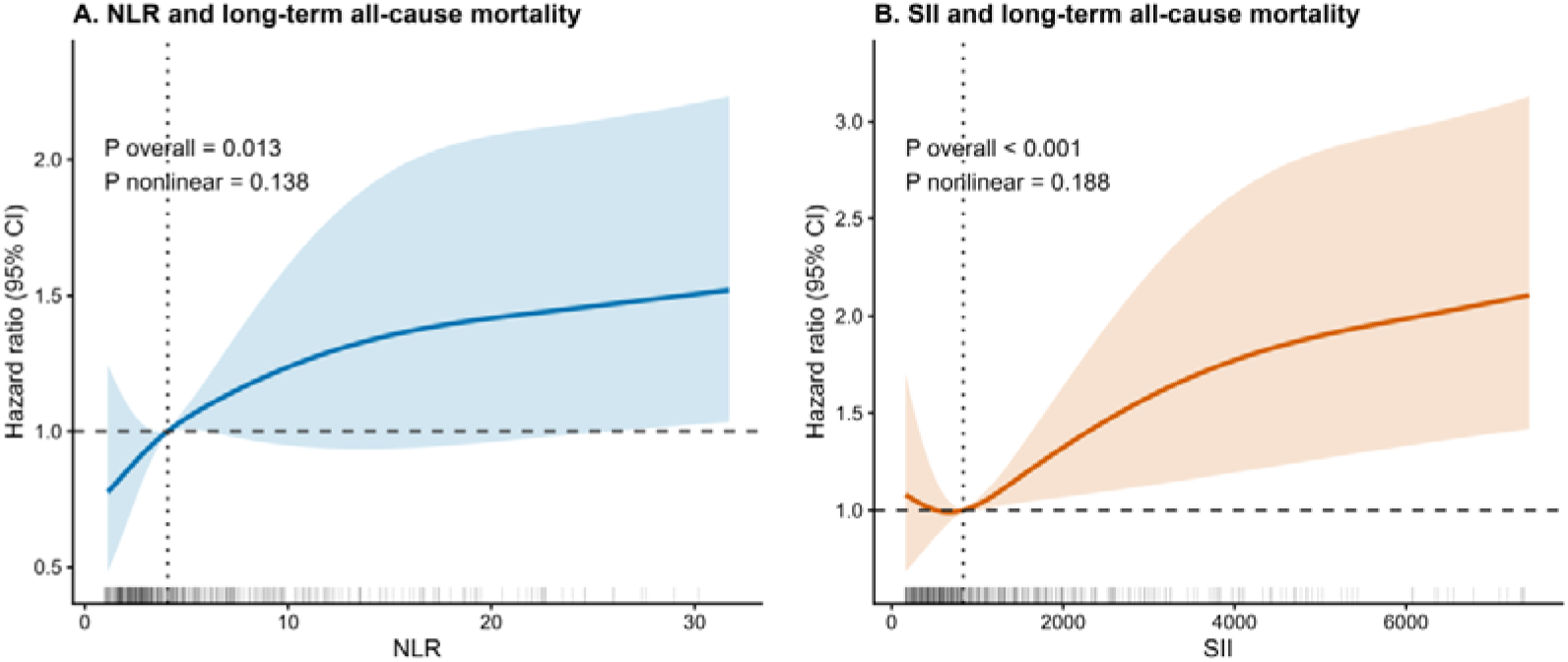
Restricted cubic spline analyses of NLR, SII, and long-term all-cause mortality in the Hong Kong cohort. Restricted cubic spline curves showing the adjusted associations of baseline NLR and SII with long-term all-cause mortality in the Hong Kong cohort. (A) NLR and long-term all-cause mortality. (B) SII and long-term all-cause mortality. Hazard ratios were estimated using Cox proportional hazards models adjusted for age, sex, renal disease, diabetes mellitus, hypertension, ischemic heart disease, and malignancy. The solid line represents the adjusted hazard ratio, and the shaded area represents the 95% confidence interval. The horizontal dashed line indicates HR = 1, and the vertical dotted line indicates the median value of the corresponding biomarker. Curves were plotted from the 2.5th to 97.5th percentiles to reduce the influence of extreme values. **Abbreviations:** NLR, neutrophil-to-lymphocyte ratio; SII, systemic immune-inflammation index; HR, hazard ratio; CI, confidence interval.

### Short-term mortality and reproducibility analyses

Associations of NLR and SII with short-term mortality in the Hong Kong cohort and mortality outcomes in the MIMIC-IV reproducibility cohort are shown in **Table 4**. In the Hong Kong cohort, 90-day all-cause mortality occurred in 33 patients. In adjusted Cox models, NLR was associated with 90-day all-cause mortality both as a continuous variable and for tertile 3 versus tertile 1. SII showed the same pattern when analyzed per 100-unit increase and for tertile 3 versus tertile 1.

**Table 4.**
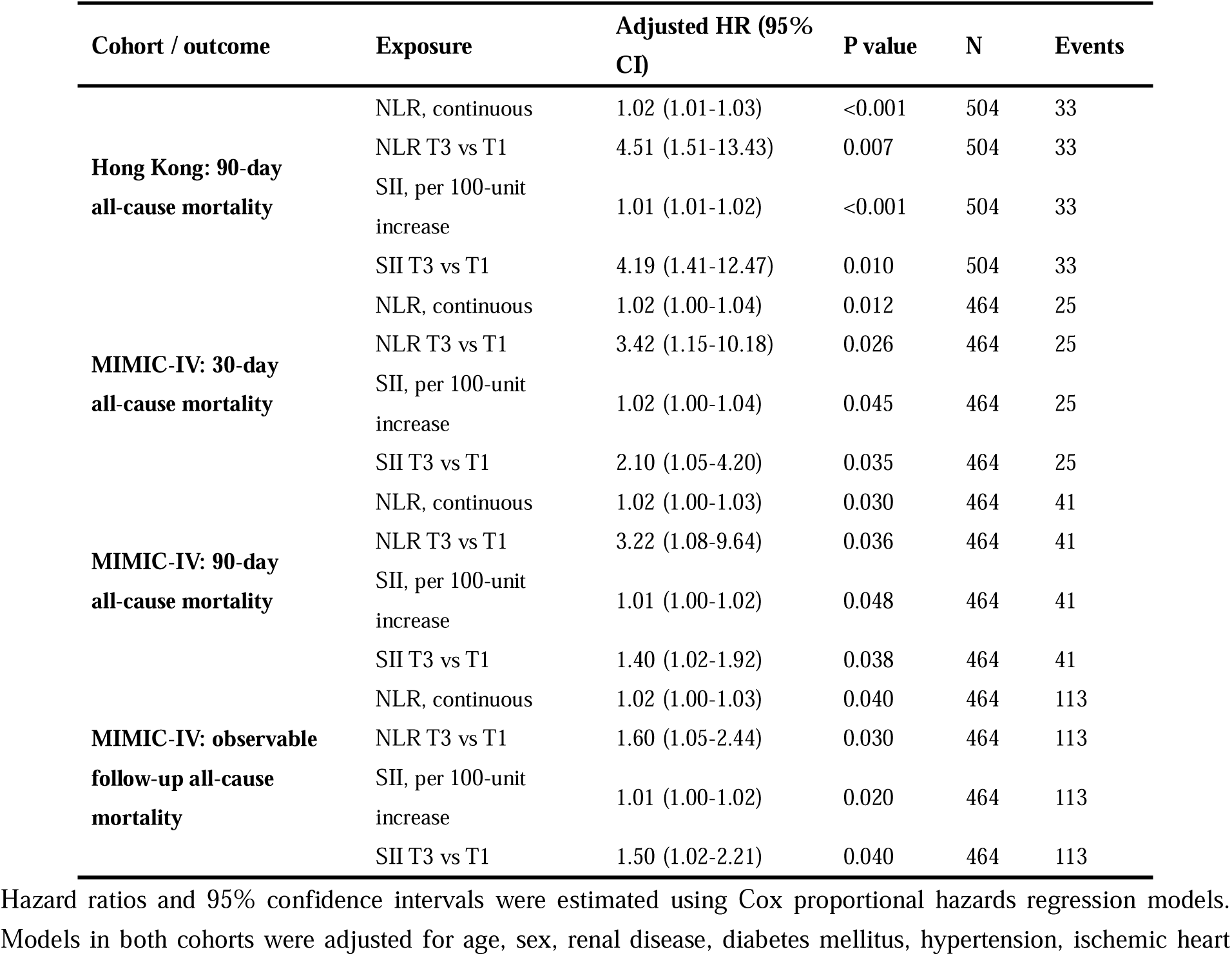
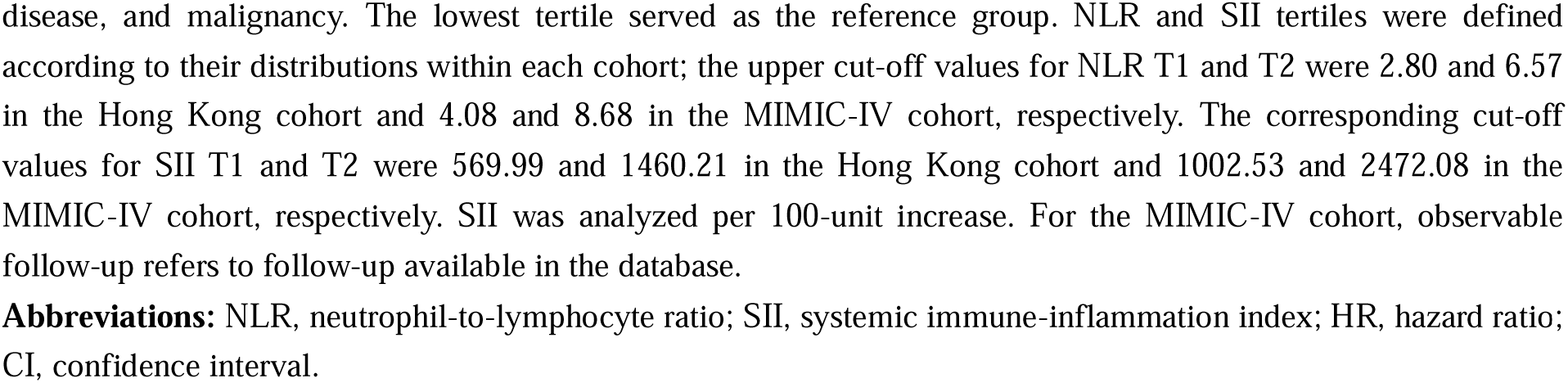
Associations of NLR and SII with short-term mortality in the Hong Kong cohort and mortality outcomes in the MIMIC-IV reproducibility cohort.

| Cohort / outcome | Exposure | Adjusted HR (95% CI) | P value | N | Events |
| --- | --- | --- | --- | --- | --- |
| **Hong Kong: 90-day all-cause mortality** | NLR, continuous | 1.02 (1.01-1.03) | <0.001 | 504 | 33 |
|  | NLR T3 vs T1 | 4.51 (1.51-13.43) | 0.007 | 504 | 33 |
|  | SII, per 100-unit increase | 1.01 (1.01-1.02) | <0.001 | 504 | 33 |
|  | SII T3 vs T1 | 4.19 (1.41-12.47) | 0.010 | 504 | 33 |
| **MIMIC-IV: 30-day all-cause mortality** | NLR, continuous | 1.02 (1.00-1.04) | 0.012 | 464 | 25 |
|  | NLR T3 vs T1 | 3.42 (1.15-10.18) | 0.026 | 464 | 25 |
|  | SII, per 100-unit increase | 1.02 (1.00-1.04) | 0.045 | 464 | 25 |
|  | SII T3 vs T1 | 2.10 (1.05-4.20) | 0.035 | 464 | 25 |
| **MIMIC-IV: 90-day all-cause mortality** | NLR, continuous | 1.02 (1.00-1.03) | 0.030 | 464 | 41 |
|  | NLR T3 vs T1 | 3.22 (1.08-9.64) | 0.036 | 464 | 41 |
|  | SII, per 100-unit increase | 1.01 (1.00-1.02) | 0.048 | 464 | 41 |
|  | SII T3 vs T1 | 1.40 (1.02-1.92) | 0.038 | 464 | 41 |
| **MIMIC-IV: observable follow-up all-cause mortality** | NLR, continuous | 1.02 (1.00-1.03) | 0.040 | 464 | 113 |
|  | NLR T3 vs T1 | 1.60 (1.05-2.44) | 0.030 | 464 | 113 |
|  | SII, per 100-unit increase | 1.01 (1.00-1.02) | 0.020 | 464 | 113 |
|  | SII T3 vs T1 | 1.50 (1.02-2.21) | 0.040 | 464 | 113 |

Hazard ratios and 95% confidence intervals were estimated using Cox proportional hazards regression models. Models in both cohorts were adjusted for age, sex, renal disease, diabetes mellitus, hypertension, ischemic heart
disease, and malignancy. The lowest tertile served as the reference group. NLR and SII tertiles were defined according to their distributions within each cohort; the upper cut-off values for NLR T1 and T2 were 2.80 and 6.57 in the Hong Kong cohort and 4.08 and 8.68 in the MIMIC-IV cohort, respectively. The corresponding cut-off values for SII T1 and T2 were 569.99 and 1460.21 in the Hong Kong cohort and 1002.53 and 2472.08 in the MIMIC-IV cohort, respectively. SII was analyzed per 100-unit increase. For the MIMIC-IV cohort, observable follow-up refers to follow-up available in the database.
**Abbreviations:** NLR, neutrophil-to-lymphocyte ratio; SII, systemic immune-inflammation index; HR, hazard ratio; CI, confidence interval.

In the MIMIC-IV cohort, 30-day all-cause mortality occurred in 25 patients and 90-day all-cause mortality occurred in 41 patients (**Table 4**). In adjusted models, NLR was associated with 30-day all-cause mortality as a continuous variable and for tertile 3 versus tertile 1. NLR was also associated with 90-day all-cause mortality and all-cause mortality during observable follow-up using both exposure parameterizations.

SII was associated with 30-day all-cause mortality, 90-day all-cause mortality, and all-cause mortality during observable follow-up in the MIMIC-IV cohort when modeled as a continuous variable. Associations for SII tertile 3 versus tertile 1 were also observed for each of these outcomes (**Table 4**).

Kaplan-Meier cumulative mortality curves in the MIMIC-IV cohort are shown in **Supplementary Figure 2** for NLR tertiles and **Supplementary Figure 3** for SII tertiles. For both indices, cumulative mortality differed across tertiles for 30-day and 90-day all-cause mortality. A forest plot summarizing the associations of the highest versus lowest tertiles of NLR and SII with all-cause mortality across cohorts and outcomes is presented in **Figure 4**. The point estimates for the highest versus lowest tertiles of both NLR and SII were above 1.0 across all evaluated outcomes. Given the limited number of early mortality events, parsimonious multivariable Cox models adjusted for age, sex, renal disease, and malignancy were further performed as sensitivity analyses (**Supplementary Table 8**). In these models, both NLR and SII remained associated with 90-day all-cause mortality in the Hong Kong cohort. In MIMIC-IV, NLR and SII were associated with 30-day, 90-day, and observable follow-up all-cause mortality.

**Figure 4.**
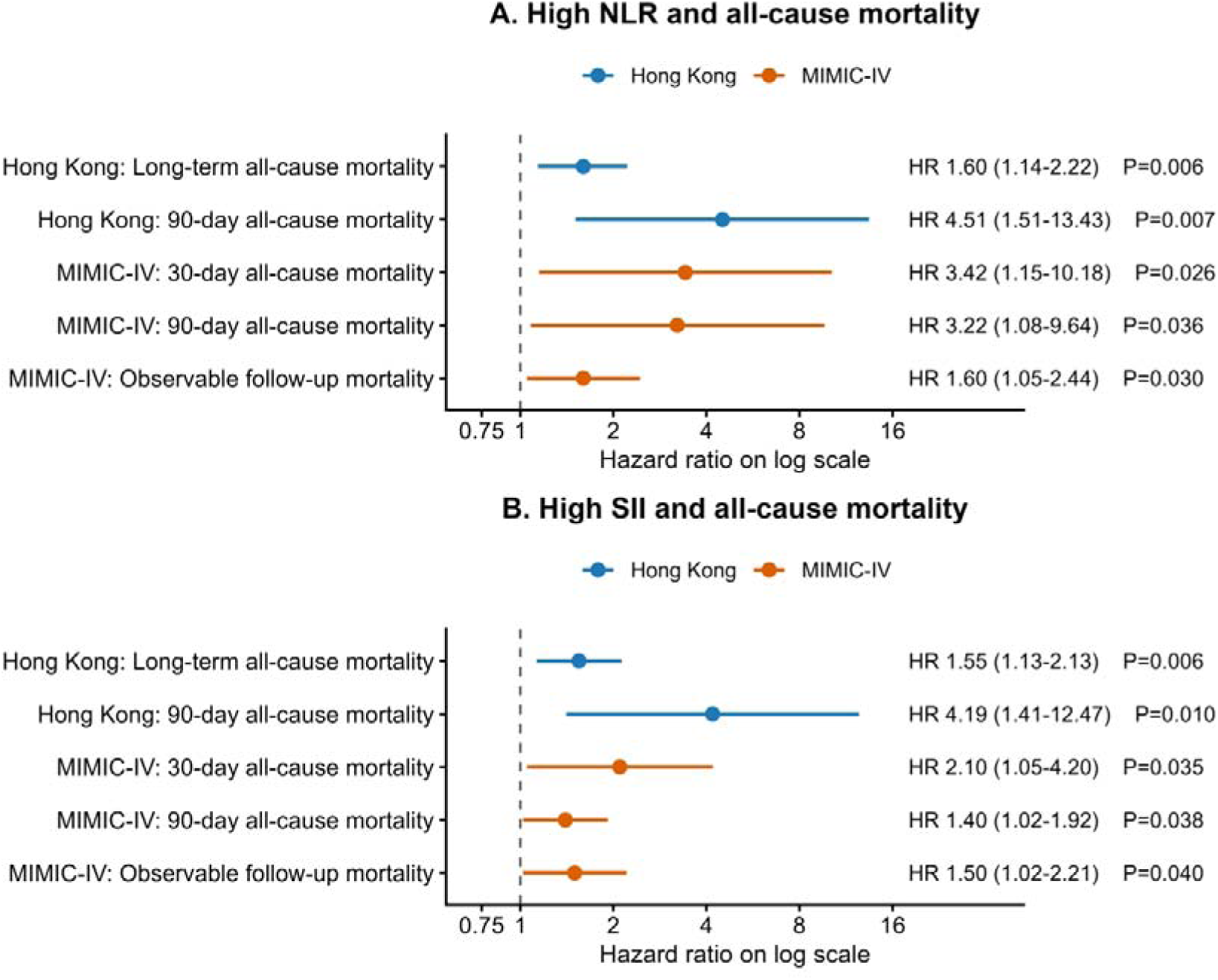
Associations of the highest versus lowest tertiles of NLR and SII with all-cause mortality across cohorts and outcomes. Forest plot showing adjusted hazard ratios for the highest versus lowest tertiles of NLR and SII across cohorts and mortality outcomes. Models were adjusted for age, sex, renal disease, diabetes mellitus, hypertension, ischemic heart disease, and malignancy. The lowest tertile served as the reference group. **Abbreviations:** NLR, neutrophil-to-lymphocyte ratio; SII, systemic immune-inflammation index; HR, hazard ratio; CI, confidence interval.

### Long-term sensitivity analyses and incremental prognostic value

Sensitivity analyses with follow-up truncated at 5 years are shown in **Supplementary Table 4**. In the fully adjusted model, NLR remained associated with 5-year all-cause mortality as a continuous variable and for tertile 3 versus tertile 1. SII was also associated with 5-year all-cause mortality using both exposure parameterizations.

Associations of individual blood cell components and composite inflammatory indices with long-term all-cause mortality are presented in **Supplementary Table 6**. White blood cell count, lymphocyte count, NLR, and SII were associated with long-term all-cause mortality when modeled per 1-SD increase. Neutrophil count and platelet count were not statistically significant.

Extreme-value sensitivity analyses are shown in **Supplementary Table 7**. After excluding patients above the 99th percentile of the corresponding biomarker, NLR and SII were associated with long-term all-cause mortality as continuous variables and for tertile 3 versus tertile 1. Similar findings were observed after winsorization at the 1st and 99th percentiles.

The incremental prognostic value of NLR and SII for long-term all-cause mortality in the Hong Kong cohort is shown in **Supplementary Table 5**. The clinical model had a Harrell’s C-index of 0.701. Addition of NLR, SII, or both indices increased the C-index and the 1-year, 3-year, and 5-year time-dependent AUCs. At 5 years, addition of NLR, SII, or both indices was associated with positive integrated discrimination improvement and continuous net reclassification improvement compared with the clinical model.

## Discussion

In this retrospective dual-cohort study of patients with pericarditis, we evaluated the prognostic value of two readily available hematologic inflammatory indices across short-term and long-term all-cause mortality outcomes. The principal findings were as follows. First, in the Hong Kong electronic health record cohort, higher baseline NLR was associated with long-term all-cause mortality after adjustment for demographic factors and major comorbidities. Second, higher NLR was also associated with 90-day all-cause mortality in the Hong Kong cohort and with 30-day, 90-day, and observable follow-up all-cause mortality in the MIMIC-IV reproducibility cohort. Third, SII showed supportive associations with mortality outcomes in both cohorts, although the magnitude of association was generally more pronounced for NLR. Fourth, dose-response and sensitivity analyses supported the consistency of these associations in the Hong Kong cohort.

Evidence on simple hematologic prognostic markers in pericarditis remains limited, particularly for mortality outcomes and cross-cohort reproducibility. Previous studies evaluating inflammatory biomarkers in pericarditis have provided important insights into disease activity, recurrence risk, and short-term complications, but most available evidence has come from small observational studies and has focused mainly on recurrence, tamponade, or inflammatory phenotypes rather than both short-term and long-term mortality^9,10,12^. The present study extends this evidence by evaluating mortality outcomes in a primary long-term electronic health record cohort and by examining reproducibility in an independent real-world database. This distinction is clinically relevant because mortality risk in patients with pericarditis may reflect not only pericardial inflammation but also systemic illness, comorbidity burden, and reduced physiological reserve^17^. Therefore, the mortality rates in this study should be interpreted as reflecting hospitalized real-world pericarditis cohorts with substantial clinical heterogeneity, rather than the prognosis of uncomplicated idiopathic or viral pericarditis.

Pericarditis is an inflammatory disorder with a broad clinical spectrum, and current risk assessment relies mainly on clinical features, suspected etiology, imaging findings, and selected inflammatory biomarkers^1–3^. In this setting, complete blood count-derived indices may provide practical information because they are inexpensive, routinely measured, and available early during hospitalization^7,18^. Rather than replacing established clinical assessment, NLR and SII may serve as adjunctive markers for identifying patients who may require closer observation, more complete clinical evaluation, or follow-up planning^1,2^.

The association between NLR and mortality may reflect the combined prognostic information conveyed by neutrophil-predominant inflammation and relative lymphopenia. NLR integrates innate inflammatory activation and adaptive immune status^6,19,20^. In patients with pericarditis, elevated NLR may not simply represent local pericardial inflammation, but may also capture a broader host-response phenotype related to systemic inflammatory activation, infection or malignancy, lymphopenia, comorbidity burden, and reduced physiological reserve^2,21^. This is clinically relevant because pericarditis may occur in the context of infection, cardiac procedures, malignancy, chronic kidney disease, or systemic autoimmune disease, and mortality in real-world cohorts may therefore reflect both pericardial disease and systemic vulnerability^2,22^. In the Hong Kong cohort, increasing NLR tertiles were accompanied by higher white blood cell and neutrophil counts and lower lymphocyte counts, whereas major clinical comorbidities included in the adjusted model did not differ significantly across tertiles. These findings suggest that NLR captured variation in inflammatory and immune status beyond the measured comorbidity profile. However, given the observational design, these associations should not be interpreted as evidence of causality.

The two cohorts differed in clinical setting, comorbidity burden, biomarker distributions, and follow-up structure. The Hong Kong cohort provided long-term electronic health record follow-up and served as the primary analysis cohort, whereas MIMIC-IV provided an independent hospital-based reproducibility cohort with shorter observable follow-up. Accordingly, absolute mortality rates, follow-up duration, and biomarker thresholds should not be directly compared between cohorts. The purpose of the MIMIC-IV analyses was not to externally validate a fixed prediction model, but to examine whether the direction of biomarker-mortality associations was reproducible in a distinct real-world setting. Despite differences in data structure and patient populations^17^, higher NLR was associated with mortality outcomes in both cohorts, supporting the cross-cohort reproducibility of the association.

SII incorporates platelet count in addition to neutrophil and lymphocyte counts and may therefore reflect inflammatory, immune, and platelet-related thromboinflammatory pathways^7,23,24^. In the present study, higher SII was associated with long-term all-cause mortality in the Hong Kong cohort and showed directionally consistent associations with short-term and observable follow-up mortality outcomes in the MIMIC-IV reproducibility analyses. However, the magnitude of association was generally less pronounced than that observed for NLR. This difference may suggest that the neutrophil-lymphocyte axis, rather than the platelet-related component, represents the dominant hematologic signal associated with mortality in this pericarditis population. This interpretation is partly supported by the component analysis, in which lymphocyte count, NLR, and SII were associated with long-term mortality, whereas platelet count alone was not statistically significant. Because NLR and SII share neutrophil and lymphocyte components, their prognostic information is likely to overlap. These findings do not necessarily indicate that SII lacks prognostic relevance; rather, they suggest that in pericarditis, the additional platelet component may add limited information once the neutrophil-lymphocyte axis is captured. Consistently, adding both indices to the clinical model did not provide clearly greater discrimination than adding either index alone. Therefore, SII may serve as a supportive inflammatory marker in patients with pericarditis, but its incremental clinical value beyond NLR, as well as the advantage of combining both indices, requires further evaluation.

Restricted cubic spline analyses showed significant overall associations of NLR and SII with long-term all-cause mortality without significant evidence of nonlinearity. These findings suggest that the associations were not confined to a single abrupt threshold. Therefore, continuous modeling quantified risk across the biomarker distribution, whereas tertile-based analyses provided clinically interpretable risk strata^25^.

Several sensitivity analyses supported the consistency of the findings. Associations of NLR and SII with 5-year all-cause mortality were consistent with the primary long-term analyses. Extreme-value analyses, including exclusion of the upper 1% and winsorization at the 1st and 99th percentiles, showed similar results, suggesting that the associations were not driven primarily by outlying biomarker values. Component analyses showed that lymphocyte count, NLR, and SII were associated with long-term mortality, whereas neutrophil and platelet counts alone were not statistically significant. In addition, parsimonious multivariable models adjusted for age, sex, renal disease, and malignancy yielded broadly consistent associations for short-term and observable follow-up mortality outcomes. These findings support the stability of the observed associations, while also highlighting the overlap among related hematologic inflammatory markers.

The clinical implications of these findings should be interpreted in the context of risk stratification rather than immediate treatment selection. NLR and SII are not disease-specific diagnostic markers and should not be used as stand-alone tools for treatment escalation. Their potential value lies in early bedside risk enrichment, particularly when interpreted alongside clinical presentation, suspected etiology, imaging findings, conventional inflammatory biomarkers, and comorbidity burden^26,27^. The modest improvement in discrimination suggests that these indices may provide risk enrichment rather than function as a stand-alone prognostic model. Because these indices may be influenced by infection, malignancy, renal dysfunction, medication exposure, and other inflammatory or systemic conditions, they should be regarded as adjunctive risk markers rather than independent decision-making biomarkers^1,7,18,28^.

This study has several strengths. The dual-cohort design allowed assessment of biomarker-outcome associations in a primary long-term electronic health record cohort and an independent reproducibility cohort^17,29^. The analysis focused on all-cause mortality outcomes, which reduced reliance on cause-specific mortality definitions that may vary across databases^30^. The use of a common covariate set in both cohorts facilitated comparability between the primary and reproducibility analyses. In addition, the study incorporated complementary analytic approaches, including categorical and continuous Cox models, Kaplan-Meier analyses, restricted cubic splines, sensitivity analyses, and incremental prognostic evaluation^25,31^.

Several limitations should be considered. First, the retrospective design is subject to residual confounding and selection bias. Although the adjusted models included major demographic and comorbidity variables available in both cohorts, unmeasured factors such as pericarditis etiology, treatment strategy, disease severity, imaging findings, and inflammatory marker trajectories may have influenced the observed associations^1^. Detailed etiologic classification was not consistently available across the two databases, although mortality risk may differ substantially among infectious, malignant, renal failure-related, post-procedural, autoimmune, and idiopathic pericarditis. Second, NLR and SII were unavailable in a proportion of eligible patients, particularly in the Hong Kong cohort, which may have introduced selection bias. Third, both cohorts consisted of hospitalized patients, and MIMIC-IV represents a highly selected hospital-based population; therefore, the findings may not be generalizable to mild outpatient or uncomplicated idiopathic or viral pericarditis. Fourth, pericarditis identification relied on clinical diagnosis and diagnostic codes in electronic health records, and misclassification cannot be fully excluded^32^. Fifth, CRP and other inflammatory or cardiac injury biomarkers were not included in the primary multivariable models because of missingness and limited cross-cohort availability. Therefore, the independence of NLR and SII relative to other inflammatory markers could not be fully assessed. Moreover, NLR and SII were measured only at baseline, and serial changes in these indices were not evaluated. Sixth, the two cohorts differed in clinical setting and follow-up structure. The Hong Kong cohort provided longer electronic health record follow-up, whereas MIMIC-IV provided shorter observable follow-up from a hospital-based database^17^. As a result, mortality rates and follow-up duration should not be directly compared across cohorts. In addition, although parsimonious models were used as sensitivity analyses to reduce the risk of overfitting, the numbers of early deaths remained limited, and the short-term mortality findings should be interpreted as exploratory and confirmed in larger cohorts. Finally, this study evaluated associations rather than causal effects, and the findings require prospective validation before NLR or SII can be incorporated into formal management algorithms for pericarditis.

Future studies should prospectively evaluate whether baseline and serial changes in NLR and SII are associated with disease activity, treatment response, and long-term prognosis in patients with pericarditis. Studies incorporating pericarditis etiology, imaging findings, CRP and other inflammatory biomarkers, treatment strategies, and longitudinal biomarker trajectories are also needed to determine whether complete blood count-derived inflammatory indices can improve clinically actionable risk stratification beyond established assessment strategies.

## Conclusions

In this retrospective dual-cohort study of hospitalized patients with pericarditis, higher baseline NLR and SII were associated with increased all-cause mortality. These associations were observed in the Hong Kong primary analysis cohort and were directionally consistent in the MIMIC-IV reproducibility cohort. NLR showed a more consistent prognostic signal than SII. These complete blood count–derived indices may serve as simple adjunctive markers for mortality risk stratification, although prospective validation is required before clinical implementation.

## Declarations

### Ethics approval and consent to participate

Ethics approval covering the source dataset and related secondary analyses was obtained from the Chinese University of Hong Kong–New Territories East Cluster Clinical Research Ethics Committee (approval numbers: 2018.309, 2019.338, 2019.361, and 2019.422). The requirement for informed consent was waived by the Ethics Committee because of the retrospective use of de-identified data and the absence of patient contact. The study was conducted in accordance with the Declaration of Helsinki. The corresponding approval records are listed in the CUHK–NTEC CREC approved-study database.

## Consent for publication

Not applicable.

## Data Availability Statement

This study used a dataset derived from a previously conducted retrospective cohort of adult patients hospitalized for pericarditis at a single tertiary centre in Hong Kong between 2005 and 2019. The dataset was generated as part of the doctoral thesis work of Dr Gary Tse at The Chinese University of Hong Kong and is hosted in ProQuest Dissertations & Theses Global (ProQuest document ID: 3252768154). Due to institutional data governance requirements and patient privacy regulations, the underlying original raw electronic health record data cannot be openly shared and are no longer kept by the authors. MIMIC-IV is a publicly accessible, deidentified database available through PhysioNet to credentialed users who complete the required training and data use agreement.

## Acknowledgments

Not applicable.

## Author Contributions

Lingyu Mi contributed to the conceptualization, methodology, formal analysis, investigation, data curation, visualization, and preparation of the original manuscript. Ishan Lakhani contributed to data curation, methodology, validation, and critical revision of the manuscript. Wing Tak Wong contributed to methodology, validation, data interpretation, and critical revision of the manuscript. Gary Tse contributed to conceptualization, methodology, resources, supervision, project administration, data interpretation, and critical revision of the manuscript. Fang Fang contributed to conceptualization, methodology, supervision, project administration, funding acquisition, data interpretation, and critical revision of the manuscript.

