## Supplemental Data 1 for "Association of Neutrophil-to-Lymphocyte Ratio and Systemic Immune-Inflammation Index With Mortality in Patients With Pericarditis: A Retrospective Dual-Cohort Study Using Two Independent Databases"

**Supplementary Appendix**

**Supplementary Table 1.** Baseline characteristics and clinical outcomes of patients with and without available NLR and SII in the Hong Kong cohort.

| **Characteristic** | **Available NLR and SII (n=504)** | **Missing NLR or SII (n=370)** | **P value** |
| --- | --- | --- | --- |
| **Demographics** |  |  |  |
| Age, years | 60.70 (50.19, 71.78) | 58.15 (49.22, 67.70) | 0.053 |
| Male sex, n (%) | 263 (52.2%) | 231 (62.4%) | 0.003 |
| **Comorbidities** |  |  |  |
| History of VT/VF, n (%) | 4 (0.8%) | 4 (1.1%) | 0.728 |
| Cerebrovascular disease, n (%) | 17 (3.4%) | 11 (3.0%) | 0.891 |
| Renal disease, n (%) | 33 (6.5%) | 15 (4.1%) | 0.147 |
| Diabetes mellitus, n (%) | 14 (2.8%) | 11 (3.0%) | 1.000 |
| Hypertension, n (%) | 97 (19.2%) | 58 (15.7%) | 0.202 |
| Acute myocardial infarction, n (%) | 11 (2.2%) | 12 (3.2%) | 0.451 |
| COPD, n (%) | 5 (1.0%) | 3 (0.8%) | 1.000 |
| Ischemic heart disease, n (%) | 55 (10.9%) | 31 (8.4%) | 0.259 |
| Peripheral vascular disease, n (%) | 6 (1.2%) | 6 (1.6%) | 0.805 |
| Malignancy, n (%) | 101 (20.0%) | 64 (17.3%) | 0.349 |
| Metastatic solid tumor, n (%) | 47 (9.3%) | 47 (12.7%) | 0.138 |
| **Laboratory indices** |  |  |  |
| White blood cell count, ×10⁹/L | 7.05 (5.50, 9.03) | 6.60 (5.95, 9.45) | 0.680 |
| Platelet count, ×10⁹/L | 210.00 (163.00, 274.25) | 238.00 (187.50, 295.00) | 0.093 |
| Creatinine, μmol/L | 85.00 (66.00, 119.00) | 80.00 (66.00, 105.00) | 0.083 |
| Glucose, mmol/L | 6.39 (5.41, 8.20) | 6.45 (5.62, 8.09) | 0.328 |
| **Outcomes and follow-up** |  |  |  |
| 90-day all-cause mortality, n (%) | 33 (6.5%) | 47 (12.7%) | 0.003 |
| Long-term all-cause mortality during follow-up, n (%) | 241 (47.8%) | 155 (41.9%) | 0.095 |
| Follow-up duration, days | 1980.00 (967.75, 3172.25) | 829.50 (265.00, 1850.75) | <0.001 |

Continuous variables are presented as median (interquartile range), and categorical variables as n (%). P values were calculated using the Wilcoxon rank-sum test for continuous variables and the chi-square test or Fisher's exact test for categorical variables, as appropriate. Patients were classified according to the availability of both NLR and SII, which were required for inclusion in the main analyses.

**Abbreviations:** NLR, neutrophil-to-lymphocyte ratio; VT/VF, ventricular tachycardia/ventricular fibrillation; COPD, chronic obstructive pulmonary disease.

**Supplementary Table 2.** Baseline characteristics and clinical outcomes of patients with and without available NLR and SII in the MIMIC-IV cohort.

| **Characteristic** | **Available NLR and SII (n=464)** | **Missing NLR or SII (n=197)** | **P value** |
| --- | --- | --- | --- |
| **Demographics** |  |  |  |
| Age, years | 59.00 (46.00, 69.00) | 60.00 (47.00, 72.00) | 0.622 |
| Male sex, n (%) | 240 (51.7%) | 134 (68.0%) | <0.001 |
| **Comorbidities** |  |  |  |
| History of VT/VF, n (%) | 22 (4.7%) | 14 (7.1%) | 0.299 |
| Cerebrovascular disease, n (%) | 29 (6.2%) | 8 (4.1%) | 0.350 |
| Renal disease, n (%) | 173 (37.3%) | 65 (33.0%) | 0.336 |
| Diabetes mellitus, n (%) | 108 (23.3%) | 54 (27.4%) | 0.302 |
| Hypertension, n (%) | 194 (41.8%) | 84 (42.6%) | 0.911 |
| Acute myocardial infarction, n (%) | 53 (11.4%) | 19 (9.6%) | 0.593 |
| COPD, n (%) | 55 (11.9%) | 29 (14.7%) | 0.376 |
| Ischemic heart disease, n (%) | 108 (23.3%) | 64 (32.5%) | 0.018 |
| Peripheral vascular disease, n (%) | 36 (7.8%) | 20 (10.2%) | 0.391 |
| Malignancy, n (%) | 90 (19.4%) | 22 (11.2%) | 0.014 |
| Metastatic solid tumor, n (%) | 42 (9.1%) | 10 (5.1%) | 0.114 |
| **Laboratory indices** |  |  |  |
| White blood cell count, ×10⁹/L | 9.90 (7.50, 13.40) | 8.90 (7.20, 12.00) | 0.040 |
| Platelet count, ×10⁹/L | 258.00 (196.75, 345.50) | 222.00 (168.50, 284.50) | <0.001 |
| Creatinine, μmol/L | 88.40 (70.72, 106.08) | 88.40 (70.72, 106.08) | 0.861 |
| Glucose, mmol/L | 6.33 (5.44, 7.56) | 6.17 (5.33, 7.44) | 0.199 |
| **Outcomes and follow-up** |  |  |  |
| 30-day all-cause mortality, n (%) | 25 (5.4%) | 6 (3.0%) | 0.271 |
| 90-day all-cause mortality, n (%) | 41 (8.8%) | 12 (6.1%) | 0.302 |
| All-cause mortality during observable follow-up, n (%) | 113 (24.4%) | 34 (17.3%) | 0.057 |
| Follow-up duration, days | 175.00 (40.00, 487.00) | 207.00 (59.00, 1267.00) | 0.329 |

Continuous variables are presented as median (interquartile range), and categorical variables as n (%). P values were calculated using the Wilcoxon rank-sum test for continuous variables and the chi-square test or Fisher's exact test for categorical variables, as appropriate. Patients were classified according to the availability of both NLR and SII, which were required for inclusion in the main analyses.

**Abbreviations:** NLR, neutrophil-to-lymphocyte ratio; VT/VF, ventricular tachycardia/ventricular fibrillation; COPD, chronic obstructive pulmonary disease.

**Supplementary Table 3.** Missingness of key variables in the Hong Kong and MIMIC-IV cohorts.

| **Variable** | **Hong Kong available, n/N** | **Hong Kong missing, n/N (%)** | **MIMIC-IV available, n/N** | **MIMIC-IV missing, n/N (%)** |
| --- | --- | --- | --- | --- |
| Age | 874/874 | 0/874 (0.0%) | 661/661 | 0/661 (0.0%) |
| Male sex | 874/874 | 0/874 (0.0%) | 661/661 | 0/661 (0.0%) |
| NLR | 504/874 | 370/874 (42.3%) | 464/661 | 197/661 (29.8%) |
| SII | 504/874 | 370/874 (42.3%) | 464/661 | 197/661 (29.8%) |
| White blood cell count | 543/874 | 331/874 (37.9%) | 646/661 | 15/661 (2.3%) |
| Platelet count | 543/874 | 331/874 (37.9%) | 647/661 | 14/661 (2.1%) |
| Creatinine | 682/874 | 192/874 (22.0%) | 646/661 | 15/661 (2.3%) |
| Glucose | 467/874 | 407/874 (46.6%) | 645/661 | 16/661 (2.4%) |
| 90-day all-cause mortality | 874/874 | 0/874 (0.0%) | 661/661 | 0/661 (0.0%) |
| All-cause mortality during cohort-specific follow-up | 874/874 | 0/874 (0.0%) | 661/661 | 0/661 (0.0%) |
| Follow-up duration | 874/874 | 0/874 (0.0%) | 661/661 | 0/661 (0.0%) |

Values are presented as available n/N and missing n/N (%). Variables not recorded or not extractable in a cohort were treated as missing for that cohort. Cohort-specific follow-up refers to long-term electronic health record follow-up in the Hong Kong cohort and observable follow-up available in the MIMIC-IV database.

**Abbreviations:** NLR, neutrophil-to-lymphocyte ratio; SII, systemic immune-inflammation index.

**Supplementary Table 4.** Sensitivity analysis of the associations of NLR and SII with 5-year all-cause mortality in the Hong Kong cohort.

|  | **Model 1** | | **Model 2** | | **Model 3** | |
| --- | --- | --- | --- | --- | --- | --- |
| **Exposure** | **HR (95% CI)** | **P value** | **HR (95% CI)** | **P value** | **HR (95% CI)** | **P value** |
| **NLR** |  |  |  |  |  |  |
| Continuous variable | 1.01 (1.00-1.02) | 0.001 | 1.01 (1.00-1.02) | 0.006 | 1.01 (1.00-1.02) | 0.002 |
| Tertiles |  | P for trend <0.001 |  | P for trend <0.001 |  | P for trend <0.001 |
| T1 | Ref. |  | Ref. |  | Ref. |  |
| T2 | 1.53 (1.02-2.28) | 0.040 | 1.41 (0.94-2.12) | 0.094 | 1.46 (0.97-2.19) | 0.068 |
| T3 | 2.20 (1.50-3.23) | <0.001 | 1.93 (1.30-2.86) | 0.001 | 2.01 (1.35-3.00) | <0.001 |
| **SII** |  |  |  |  |  |  |
| Per 100-unit increase | 1.01 (1.00-1.01) | <0.001 | 1.01 (1.00-1.01) | <0.001 | 1.01 (1.00-1.01) | <0.001 |
| Tertiles |  | P for trend <0.001 |  | P for trend 0.001 |  | P for trend 0.001 |
| T1 | Ref. |  | Ref. |  | Ref. |  |
| T2 | 1.52 (1.02-2.25) | 0.039 | 1.30 (0.87-1.94) | 0.203 | 1.26 (0.84-1.90) | 0.258 |
| T3 | 2.01 (1.37-2.95) | <0.001 | 1.89 (1.28-2.78) | 0.001 | 1.89 (1.28-2.80) | 0.001 |

Hazard ratios and 95% confidence intervals were estimated using Cox proportional hazards regression models with follow-up truncated at 5 years; deaths occurring after 5 years were censored at 5 years. The lowest tertile served as the reference group. Model 1 was unadjusted. Model 2 was adjusted for age and sex. Model 3 was adjusted for age, sex, renal disease, diabetes mellitus, hypertension, ischemic heart disease, and malignancy. Patients were divided into tertiles according to the distributions of baseline NLR and SII in the Hong Kong cohort; the upper cut-off values for NLR T1 and T2 were 2.80 and 6.57, respectively, and the corresponding cut-off values for SII were 569.99 and 1460.21, respectively.

**Abbreviations:** NLR, neutrophil-to-lymphocyte ratio; SII, systemic immune-inflammation index; HR, hazard ratio; CI, confidence interval.

**Supplementary Table 5.** Incremental prognostic value of NLR and SII for long-term all-cause mortality in the Hong Kong cohort.

| **Model** | **Harrell's C-index (95% CI)** | **1-year AUC** | **3-year AUC** | **5-year AUC** | **5-year IDI (95% CI)** | **P for IDI** | **5-year continuous NRI (95% CI)** | **P for NRI** |
| --- | --- | --- | --- | --- | --- | --- | --- | --- |
| **Clinical model** | 0.701 (0.663-0.739) | 0.709 | 0.703 | 0.736 | Ref. |  | Ref. |  |
| **Clinical model + NLR** | 0.708 (0.671-0.745) | 0.725 | 0.715 | 0.745 | 0.012 (0.003-0.021) | 0.008 | 0.227 (0.065-0.412) | 0.007 |
| **Clinical model + SII** | 0.713 (0.676-0.750) | 0.736 | 0.720 | 0.751 | 0.015 (0.006-0.024) | 0.001 | 0.248 (0.090-0.373) | 0.013 |
| **Clinical model + NLR + SII** | 0.711 (0.674-0.749) | 0.734 | 0.716 | 0.751 | 0.018 (0.009-0.027) | ＜0.001 | 0.234 (0.094-0.434) | 0.013 |

Harrell's C-index and time-dependent areas under the receiver operating characteristic curves were calculated for long-term all-cause mortality. Incremental discrimination improvement and continuous net reclassification improvement were calculated at 5 years, using the clinical model as the reference model. The clinical model included age, sex, renal disease, diabetes mellitus, hypertension, ischemic heart disease, and malignancy. The clinical model was used as the reference model for IDI and NRI calculations. SII was modeled per 100-unit increase.

**Abbreviations:** NLR, neutrophil-to-lymphocyte ratio; SII, systemic immune-inflammation index; IDI, integrated discrimination improvement; NRI, net reclassification improvement; AUC, area under the curve.

**Supplementary Table 6.** Associations of blood cell components and composite inflammatory indices with long-term all-cause mortality in the Hong Kong cohort.

| **Exposure** | **Scale** | **Adjusted HR (95% CI)** | **P value** | **N** | **Events** |
| --- | --- | --- | --- | --- | --- |
| **White blood cell count** | Per 1-SD increase | 1.35 (1.20-1.53) | <0.001 | 504 | 241 |
| **Neutrophil count** | Per 1-SD increase | 1.13 (0.99-1.29) | 0.061 | 504 | 241 |
| **Lymphocyte count** | Per 1-SD increase | 0.71 (0.60-0.85) | <0.001 | 504 | 241 |
| **Platelet count** | Per 1-SD increase | 1.11 (0.98-1.27) | 0.097 | 504 | 241 |
| **NLR** | Per 1-SD increase | 1.14 (1.04-1.25) | 0.004 | 504 | 241 |
| **SII** | Per 1-SD increase | 1.21 (1.10-1.33) | <0.001 | 504 | 241 |

Hazard ratios and 95% confidence intervals were estimated using Cox proportional hazards regression models. All exposures were standardized, and hazard ratios are presented per 1-SD increase. Models were adjusted for age, sex, renal disease, diabetes mellitus, hypertension, ischemic heart disease, and malignancy.
**Abbreviations:** NLR, neutrophil-to-lymphocyte ratio; SII, systemic immune-inflammation index; HR, hazard ratio; CI, confidence interval; SD, standard deviation.

**Supplementary Table 7.** Extreme-value sensitivity analyses for the associations of NLR and SII with long-term all-cause mortality in the Hong Kong cohort.

| **Scenario** | **Exposure** | **HR (95% CI)** | **P value** | **N** | **Events** |
| --- | --- | --- | --- | --- | --- |
| **Excluding upper 1%** | NLR, continuous | 1.03 (1.02-1.05) | <0.001 | 498 | 237 |
| **Excluding upper 1%** | NLR T3 vs T1 | 1.56 (1.12-2.19) | 0.009 | 498 | 237 |
| **Excluding upper 1%** | SII, per 100-unit increase | 1.01 (1.01-1.02) | <0.001 | 498 | 236 |
| **Excluding upper 1%** | SII T3 vs T1 | 1.57 (1.14-2.17) | 0.006 | 498 | 236 |
| **Winsorization at 1st and 99th percentiles** | NLR, continuous | 1.03 (1.01-1.04) | <0.001 | 504 | 241 |
| **Winsorization at 1st and 99th percentiles** | NLR T3 vs T1 | 1.58 (1.13-2.21) | 0.007 | 504 | 241 |
| **Winsorization at 1st and 99th percentiles** | SII, per 100-unit increase | 1.01 (1.01-1.02) | <0.001 | 504 | 241 |
| **Winsorization at 1st and 99th percentiles** | SII T3 vs T1 | 1.61 (1.17-2.22) | 0.004 | 504 | 241 |

Hazard ratios and 95% confidence intervals were estimated using Cox proportional hazards regression models. Extreme-value sensitivity analyses were performed by excluding patients above the 99th percentile of the corresponding biomarker and by winsorizing biomarker values at the 1st and 99th percentiles. Models were adjusted for age, sex, renal disease, diabetes mellitus, hypertension, ischemic heart disease, and malignancy. Sample sizes varied across analyses because extreme-value handling was performed separately for each biomarker. NLR tertiles were defined using upper cut-off values of 2.80 and 6.57 for T1 and T2, respectively. SII tertiles were defined using upper cut-off values of 569.99 and 1460.21 for T1 and T2, respectively.

**Abbreviations:** NLR, neutrophil-to-lymphocyte ratio; SII, systemic immune-inflammation index; HR, hazard ratio; CI, confidence interval.

**Supplementary Table 8.** Parsimonious multivariable Cox models for short-term and observable follow-up mortality outcomes.

| **Cohort / outcome** | **Exposure** | **Parsimonious adjusted HR (95% CI)** | **P value** | **N** | **Events** |
| --- | --- | --- | --- | --- | --- |
| **Hong Kong: 90-day all-cause mortality** | NLR, continuous | 1.02 (1.01-1.03) | <0.001 | 504 | 33 |
| **Hong Kong: 90-day all-cause mortality** | NLR T3 vs T1 | 5.09 (1.71-15.14) | 0.003 | 504 | 33 |
| **Hong Kong: 90-day all-cause mortality** | SII, per 100-unit increase | 1.01 (1.01-1.02) | <0.001 | 504 | 33 |
| **Hong Kong: 90-day all-cause mortality** | SII T3 vs T1 | 4.62 (1.56-13.71) | 0.006 | 504 | 33 |
| **MIMIC-IV: 30-day all-cause mortality** | NLR, continuous | 1.02 (1.00-1.04) | 0.012 | 464 | 25 |
| **MIMIC-IV: 30-day all-cause mortality** | NLR T3 vs T1 | 3.58 (1.12-11.45) | 0.031 | 464 | 25 |
| **MIMIC-IV: 30-day all-cause mortality** | SII, per 100-unit increase | 1.02 (1.00-1.03) | 0.042 | 464 | 25 |
| **MIMIC-IV: 30-day all-cause mortality** | SII T3 vs T1 | 1.95 (1.08-3.51) | 0.027 | 464 | 25 |
| **MIMIC-IV: 90-day all-cause mortality** | NLR, continuous | 1.02 (1.00-1.04) | 0.024 | 464 | 41 |
| **MIMIC-IV: 90-day all-cause mortality** | NLR T3 vs T1 | 3.19 (1.07-9.54) | 0.038 | 464 | 41 |
| **MIMIC-IV: 90-day all-cause mortality** | SII, per 100-unit increase | 1.01 (1.00-1.02) | 0.049 | 464 | 41 |
| **MIMIC-IV: 90-day all-cause mortality** | SII T3 vs T1 | 1.78 (1.12-2.83) | 0.015 | 464 | 41 |
| **MIMIC-IV: observable follow-up all-cause mortality** | NLR, continuous | 1.02 (1.00-1.03) | 0.038 | 464 | 113 |
| **MIMIC-IV: observable follow-up all-cause mortality** | NLR T3 vs T1 | 1.52 (1.08-2.14) | 0.016 | 464 | 113 |
| **MIMIC-IV: observable follow-up all-cause mortality** | SII, per 100-unit increase | 1.01 (1.00-1.02) | 0.044 | 464 | 113 |
| **MIMIC-IV: observable follow-up all-cause mortality** | SII T3 vs T1 | 1.33 (1.02-1.73) | 0.035 | 464 | 113 |

Hazard ratios and 95% confidence intervals were estimated using Cox proportional hazards regression models. Models were adjusted for age, sex, renal disease, and malignancy. The lowest tertile served as the reference group. NLR and SII tertiles were defined according to their distributions within each cohort. SII was analyzed per 100-unit increase. For 30-day and 90-day mortality analyses, follow-up was administratively censored at 30 and 90 days, respectively. Observable follow-up refers to follow-up available in the MIMIC-IV database.

**Abbreviations:** NLR, neutrophil-to-lymphocyte ratio; SII, systemic immune-inflammation index; HR, hazard ratio; CI, confidence interval.

**Supplementary Figure 1.** Kaplan-Meier cumulative mortality curves according to SII tertiles in the Hong Kong cohort.

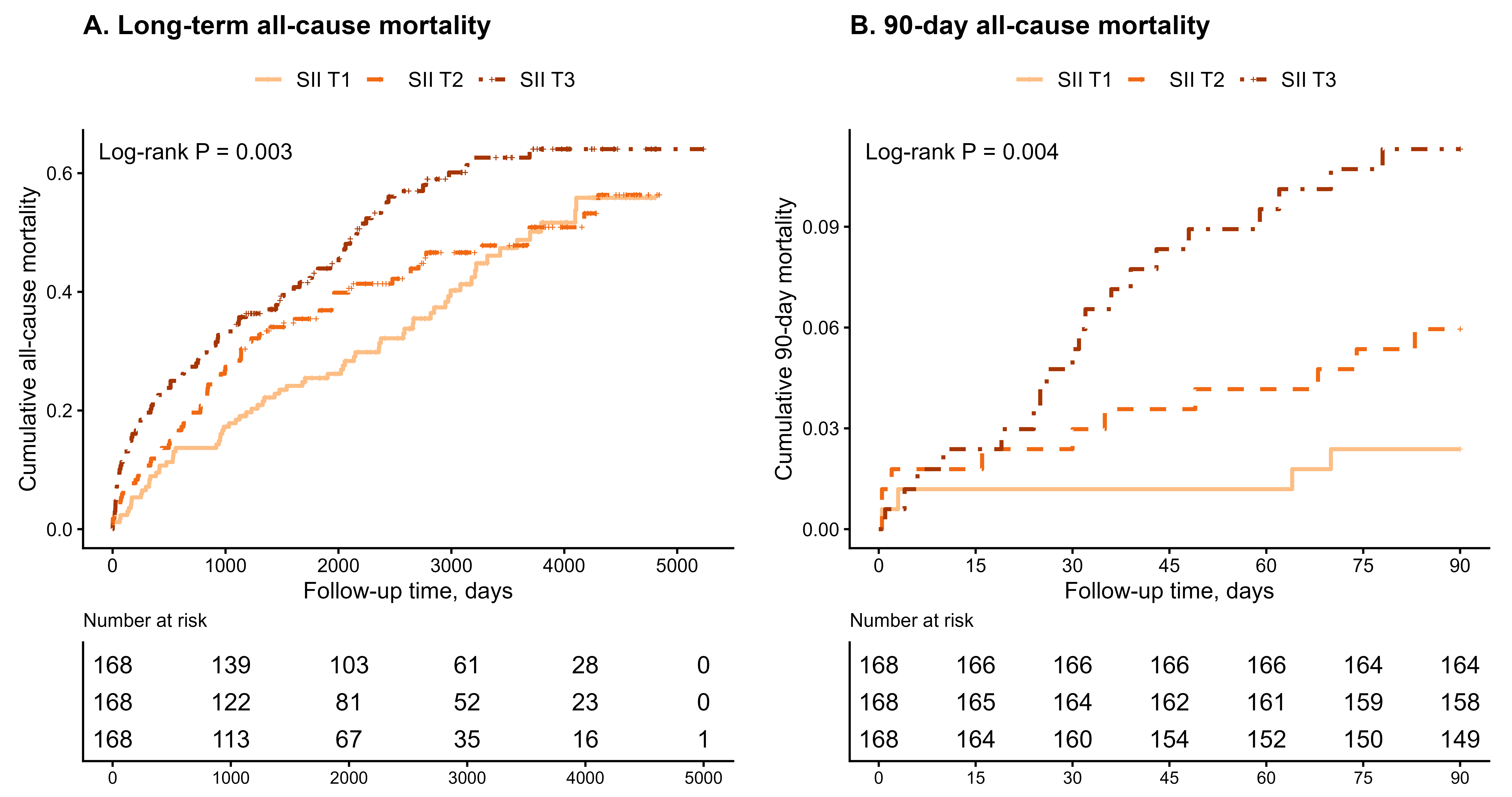

Kaplan-Meier cumulative mortality curves for patients stratified by baseline SII tertiles in the Hong Kong cohort. (A) Long-term all-cause mortality. (B) 90-day all-cause mortality. Patients were divided into SII tertiles according to the distribution of baseline SII; the upper cut-off values for SII T1 and T2 were 569.99 and 1460.21, respectively. Log-rank tests were used to compare cumulative mortality across SII tertiles.

**Abbreviations:** SII, systemic immune-inflammation index.

**Supplementary Figure 2.** Kaplan-Meier cumulative mortality curves according to NLR tertiles in the MIMIC-IV cohort.

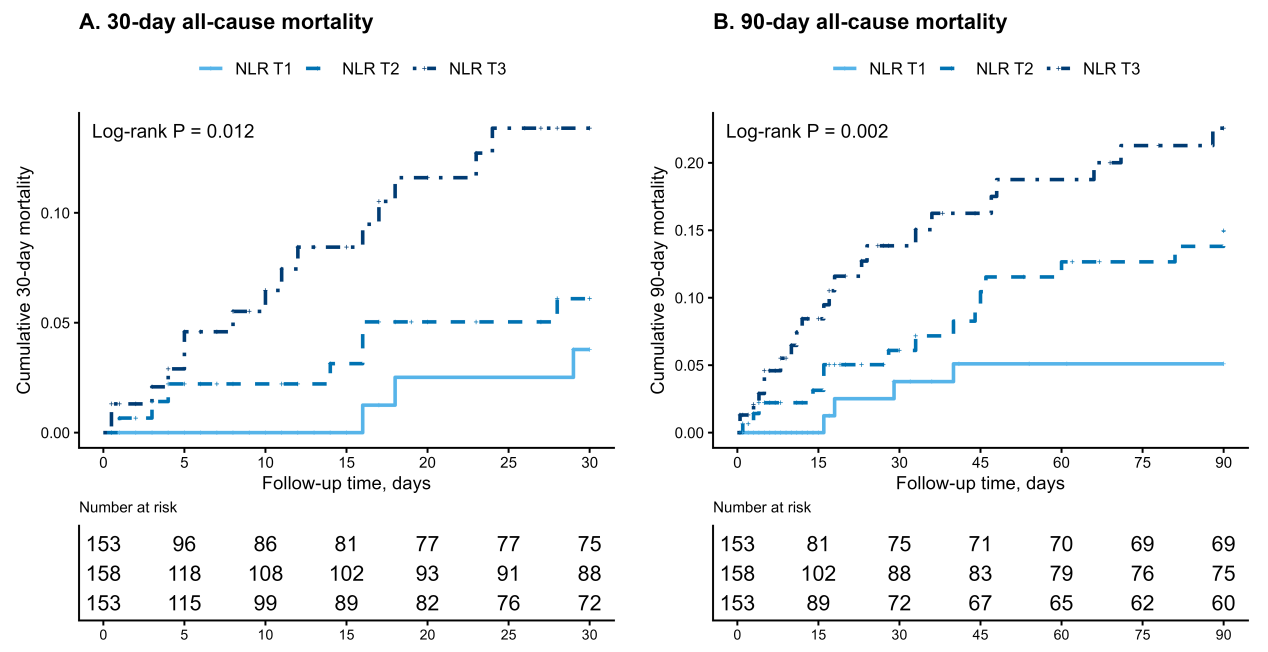

Kaplan-Meier cumulative mortality curves for patients stratified by baseline NLR tertiles in the MIMIC-IV cohort. (A) 30-day all-cause mortality. (B) 90-day all-cause mortality. Patients were divided into NLR tertiles according to the distribution of baseline NLR in the MIMIC-IV cohort; the upper cut-off values for NLR T1 and T2 were 4.08 and 8.68, respectively. Log-rank tests were used to compare cumulative mortality across NLR tertiles.
**Abbreviations:** NLR, neutrophil-to-lymphocyte ratio.

**Supplementary Figure 3.** Kaplan-Meier cumulative mortality curves according to SII tertiles in the MIMIC-IV cohort.

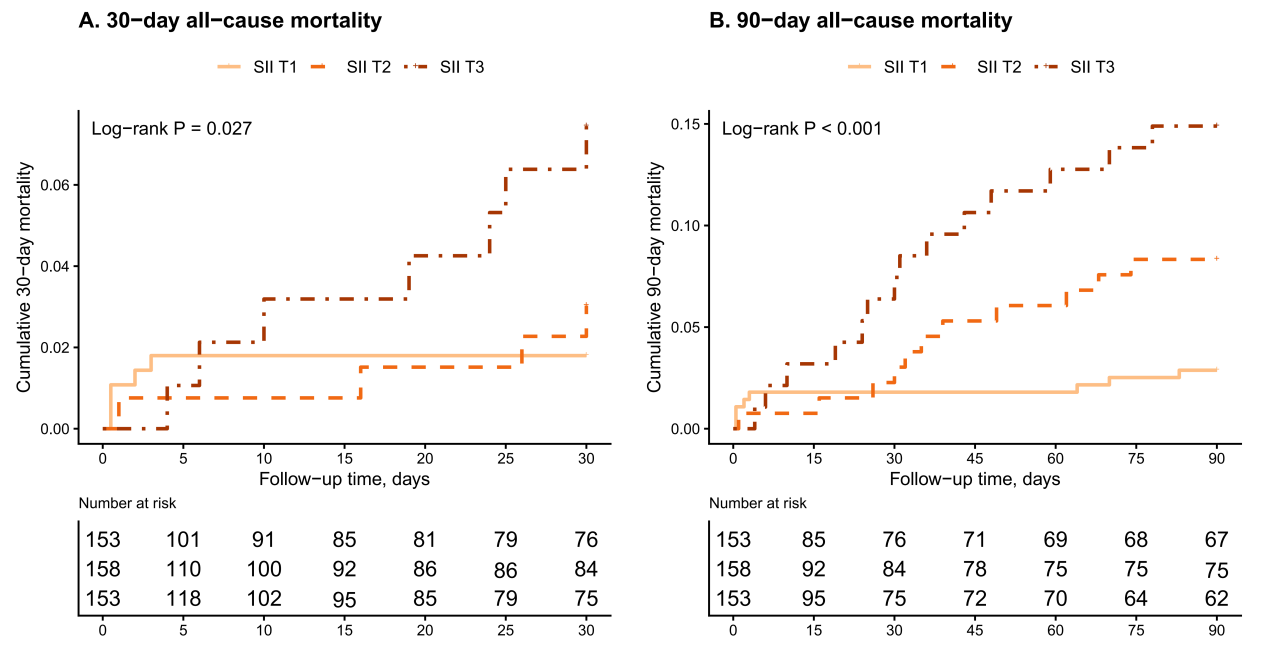

Kaplan-Meier cumulative mortality curves for patients stratified by baseline SII tertiles in the MIMIC-IV cohort. (A) 30-day all-cause mortality. (B) 90-day all-cause mortality. Patients were divided into SII tertiles according to the distribution of baseline SII in the MIMIC-IV cohort; the upper cut-off values for SII T1 and T2 were 1002.53 and 2472.08, respectively. Log-rank tests were used to compare cumulative mortality across SII tertiles.
**Abbreviations:** SII, systemic immune-inflammation index.

**Graphical Abstract**

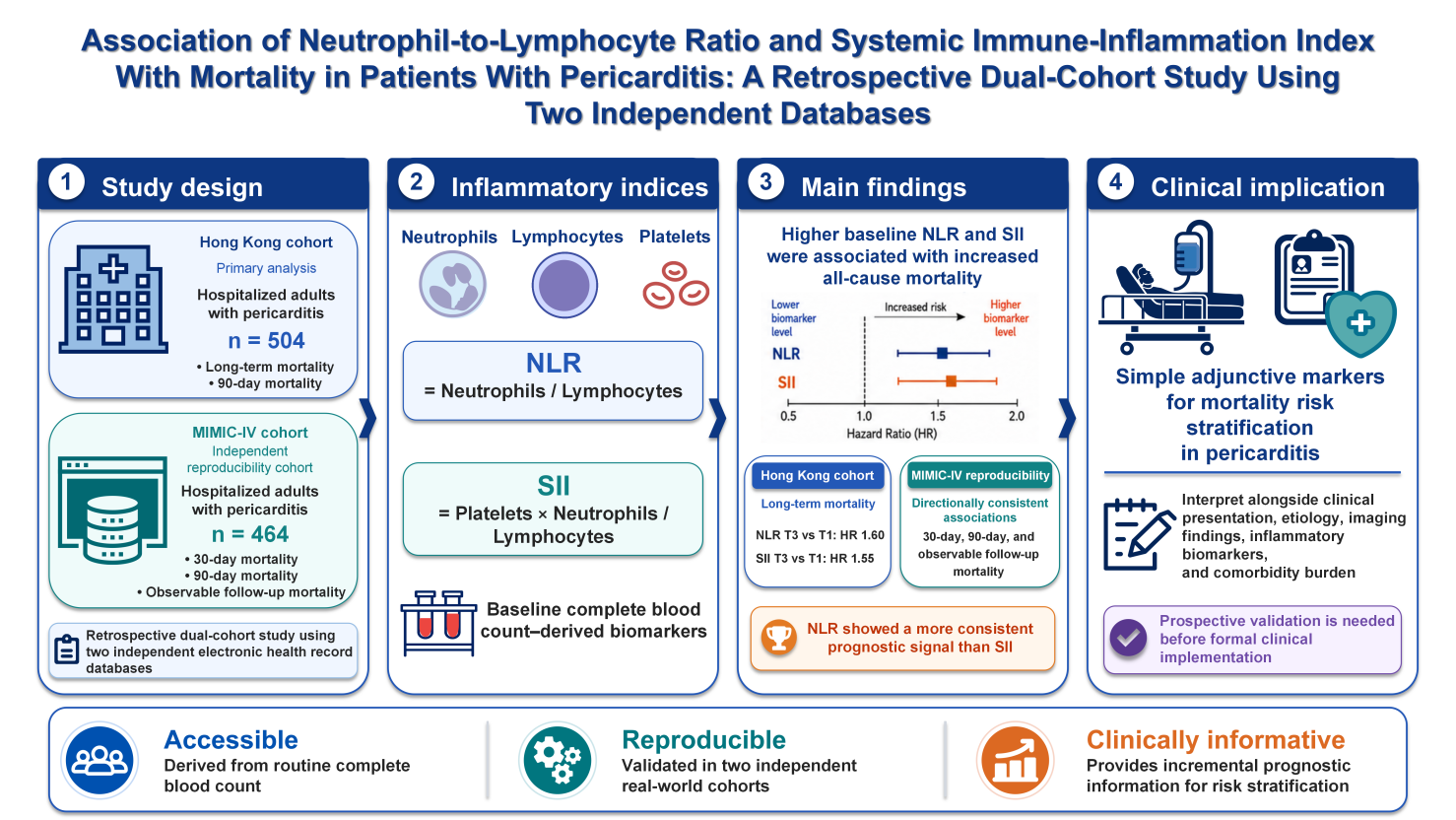
